# Data-driven estimation of the onset of heat-related mortality in five Indian cities

**DOI:** 10.64898/2026.11.08.26367540

**Authors:** Mónica M. Robles Fontán, Bhargav Krishna, Annanya Mahajan, Robert D. Meade, Abhiyant Tiwari, Satchit Balsari, Caroline Buckee, Rafael A. Irizarry

**Author notes:** **Corresponding author** Rafael A. Irizarry, PhD, Department of Data Science, Dana-Farber Cancer Institute CLSB 11007, 450 Brookline Ave, Boston, MA 02215.

## Abstract

**Background:** Extreme heat is an increasingly pressing public health threat, particularly in South Asia. Yet the temperatures at which mortality risk begins to increase in Indian cities remain poorly characterized. Operational heat wave definitions are based on meteorological criteria and may not reflect epidemiologically relevant temperature thresholds. Widely used distributed lag non-linear models are designed to flexibly estimate expected mortality as a function of temperature, but do not directly target the onset of excess risk from heat, a quantity central to early warning and planning.

**Methods:** We conducted a multi-city time-series study of daily mortality and temperature data in Ahmedabad (2001–2010), Chennai (2012–2017), Delhi (2011–2015), Kolkata (2011–2018), and Mumbai (2011–2015). We developed a data-driven structured modeling framework that permits likelihood-based inference for city-specific temperatures at which heat-related mortality first exceeds seasonal background levels, a parameter that is challenging to estimate and perform statistical inference from short and noisy series. Using these health-based thresholds, we also estimated heat-associated excess deaths.

**Results:** In all five cities, the onset of excess mortality occurred at temperatures substantially lower than those implied by current operational heat wave definitions. The proposed framework yielded stable and interpretable threshold estimates and confidence intervals across heterogeneous climates and time-series lengths and identified a larger burden of heat-associated mortality than would be captured using standard meteorological criteria.

**Conclusions:** Data-driven estimation of mortality-based heat thresholds provides epidemiologically relevant targets for early warning systems and heat action plans. Reliance on meteorological definitions alone may delay intervention and underestimate population risk in rapidly warming regions.

**Key messages:**

- We estimated the temperature at which mortality risk begins to increase above seasonal background levels in warm-climate settings and compared these estimates with current operational heat wave definitions in five Indian cities.
- Using a structured modeling approach motivated by observed patterns in the data, we found that mortality risk began to increase at temperatures below current operational heat wave thresholds, with higher-intensity heat events associated with a larger share of excess mortality.
- These findings suggest that epidemiologically derived thresholds may complement existing meteorological definitions in characterizing heat-related risk and setting triggers for action plans and warning systems.

## Introduction

Extreme heat events are among the most pressing public health challenges of the twenty-first century, particularly due to projected increases in their frequency, duration, and intensity (5). South Asia, notably India, faces rising risks from extreme heat, as seen in severe episodes in 2010, 2015, and 2022, which led to thousands of excess deaths (4; 7; 16). A ten-city study across varied agroclimatological zones in India reported a 14.7% increase in daily all-cause mortality associated with recorded temperatures above the 97th percentile for two or more consecutive days (6).

A key challenge in the study of the health effects of heatwaves is the varying criteria used to define heatwaves: absolute or relative daily maximum temperature thresholds exceeding a predetermined duration (12). The India Meteorological Department (IMD) uses climatological thresholds, including daily maximum temperatures above 40 °C in inland cities or significant (4.5–6.4 °C) departures from historical temperature trends for several consecutive days (18). These definitions serve operational forecasting but may not fully capture population health effects (21). Research shows mortality-based heat thresholds in Europe, North America, and East Asia are often lower than such operational standards (13; 19; 8; 23). Indian evidence agrees: the ten-city mortality study found significant mortality risk accumulates well before heat events qualify under IMD’s absolute temperature criteria (6). Health-based thresholds may therefore inform timely warnings and help institutions respond to heat-associated health impacts.

Contemporary epidemiological analyses use Minimum Mortality Temperature (MMT), defined as the temperature at which mortality risk is lowest, as a core metric for heat-related mortality thresholds and adaptation tracking (9; 36). Distributed lag non-linear models (DLNMs) relate daily mortality to temperatures on the same and preceding days, adjusting for seasonal trends. The MMT is derived from the cumulative exposure–response curve (estimated from a DLNM) by aggregating lagged effects (10), corresponding to a counterfactual of constant temperature over consecutive days rather than the observed lag distribution.

MMT research has motivated mortality-based heatwave definitions and yielded U- or J-shaped temperature–mortality profiles informative for policy (22). In persistently warm climates, however, relatively constant risk across wide temperature ranges, sharp increases only at high temperatures, and the need to collapse multi-lag relationships into single-temperature summaries make MMTs difficult to estimate and potentially unrepresentative of typical exposure patterns. Flexible DLNMs may also capture residual seasonal variation not fully removed by the time trend, distorting the estimated curve and producing an apparent cold effect when truly cold days are rare.

These limitations are acute in India. We analyze mortality data for Ahmedabad, Delhi, Chennai, Kolkata, and Mumbai, five of India’s seven most populous cities, over periods of 4 to 10 years depending on data availability. These tropical and subtropical cities experience predominantly warm to very hot conditions with few cold days, where flexible models may produce noisy temperature–mortality curves, making MMT difficult to estimate and potentially ill-defined.

Here, we introduce the *heat-risk onset temperature* (HOT): the temperature at which the mortality risk curve begins to rise above baseline, directly targeting the onset of excess risk for early warning in warm-climate settings. Estimating HOT poses two challenges: residual seasonal variation can distort the temperature–mortality relationship, and conventional approaches often require extensive lag structures inconsistent with the short duration of heat effects on mortality (33; 20). To address these limitations, we propose a structured model in which the temperature–mortality association is constant up to an unknown threshold and increases thereafter. By focusing on a single, well-defined exposure, HOT is directly interpretable, as it does not require aggregating temperature effects across multiple days and corresponds to observable temperature conditions. This approach reduces sensitivity to residual seasonal confounding and enables stable identification of the temperature at which risk begins to rise.

Threshold-based models have been proposed and applied in earlier temperature–mortality studies (3; 14; 25; 19). Although they have largely been replaced by more flexible approaches such as DLNMs, their simpler structure can be advantageous in settings with short and noisy time series, such as those considered here. Our approach retains the core modeling strategy underlying DLNMs, separating smooth seasonal and long-term variation from temperature-associated risk, but replaces the flexible temperature–mortality function with a structured threshold-based parameterization.

Establishing epidemiologically grounded temperature thresholds addresses an urgent governance gap with direct impact on population health in India. Heat Action Plans (HAPs), the primary governmental heatwave response framework, rely on IMD thresholds as triggers for coordinated emergency response measures. HAPs lack local, evidence-based threshold calibration, potentially delaying protective measures during heatwaves (30).The generation of more granular, health-based thresholds, as is the case with the HOT estimates in this study, can directly inform the recalibration of HAP trigger thresholds (29; 2).

## 1. Methods

### 1.1 Data

We obtained city-level daily mortality counts from municipal health departments or vital registration systems. Due to variation in record-keeping practices and data access, the study period differed by city: Ahmedabad (2001–2010), Chennai (2012–2017), Delhi (2011–2015), Kolkata (2011–2018), and Mumbai (2011–2015). Based on information from news reports, we removed periods coinciding with major non-heat-related shocks (such as epidemics, floods, or earthquakes) to avoid confounding with heat (Supplementary Table S1). Death registration completeness in the urban areas of the states/union territories containing these five cities is well over 90%, supporting confidence in the representativeness of the mortality data used in this study (28).

We used daily maximum temperature from the India Meteorological Department’s 1º ! 1º gridded dataset (1951–2018), selecting for each city the grid cell closest to the urban core with continuous coverage (Supplementary Figure S1). Maximum rather than minimum temperature was used as it better explains heat-wave-associated mortality (13).

### 1.2 Motivation

We applied a distributed lag non-linear model (DLNM) framework to assess its suitability in our setting: short time series from warm climates with limited cold exposure. Following standard practice, long-term and seasonal trends were modeled using natural cubic splines, with the number of degrees of freedom per year selected based on exploratory analysis (three for Ahmedabad, Chennai, Delhi, and Kolkata, and four for Mumbai). The temperature-mortality association was modeled using a distributed lag structure, with lag-specific effects parameterized by natural cubic splines (four equally spaced knots in the observed temperature distribution) and summarized as a cumulative effect across lags. The resulting curves showed sharp risk increases at high temperatures and mild increases at low temperatures (Figure 1). Furthermore note that for Chennai and Mumbai the curves moves up and down in the 25-30ºC range, challenging epidemiological expectations. In addition, for Ahmedabad and Kolkata, the relative risk estimate exceeds 1.2in the 20-25ºC range, which are mild/comfortable conditions.

**Figure 1.**
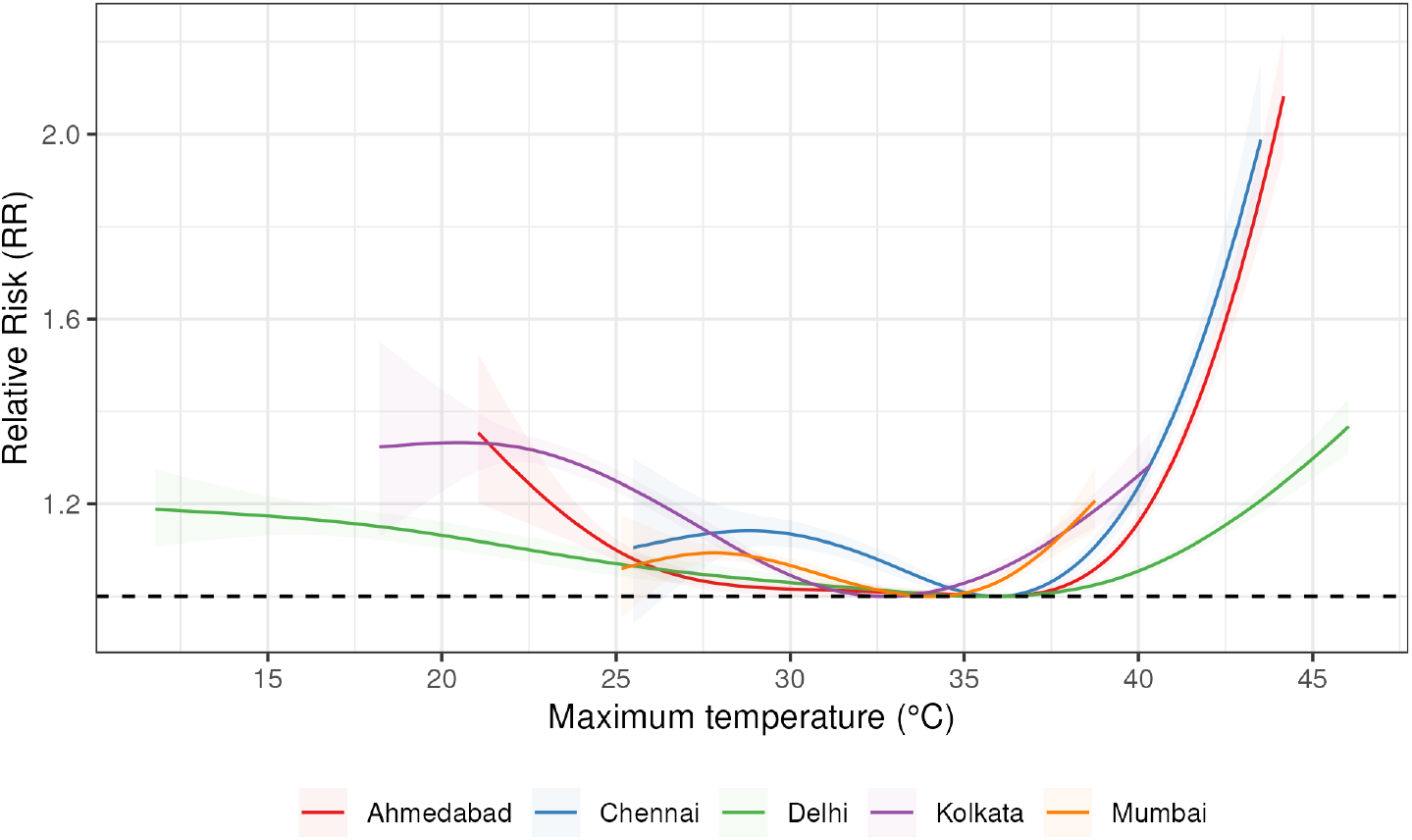
Estimated temperature-mortality curves obtained from DLNMs fitted separately for each of the five Indian cities. Following standard practice, long-term and seasonal trends were modeled using natural cubic splines, with the number of degrees of freedom per year selected based on exploratory analysis (three for Ahmedabad, Chennai, Delhi, and Kolkata, and four for Mumbai). The temperature-mortality association was modeled using a distributed lag structure, with lag-specific effects parameterized by natural cubic splines (four equally spaced knots in the observed temperature distribution) and summarized as a cumulative effect across lags.

To assess whether the estimated temperature–mortality curves reflect underlying structure or overfitting by the DLNM, we examined residuals from a model that adjusted for long-term and seasonal patterns but excluded the temperature effect. The residuals do not support a full U- or J-shaped relationship. Instead, they are consistent with a simpler pattern: mortality remains approximately constant across most temperatures and increases only beyond a high-temperature threshold, with a smooth, accelerating rise at higher temperatures (Supplementary Figure S2).

We examined the structure of temperature across lags and its relationship with mortality to better understand the role of the lag structure. A two-dimensional smoother of residual mortality against temperatures at lags *t* and *t →* 1 showed little distinct variation along either dimension when conditioning on the other, consistent with strong collinearity across lags up to 7 days (Supplementary Figures S3, S4–S8). To identify the most informative lag, we fit loess smoothers to mortality residuals at each lag and compared residual error; model fit with lag 1 consistently explained more variability than lag 0 and lags 2–7 (Supplementary Figures . After accounting for lag 1 temperature, residuals showedno systematic association with remaining lags (Supplementary Figures S15–S19), confirming that the model fit with lag 1 captures most of the relevant temperature signal.

We also examined the added explanatory power of humidity on all-cause mortality after accounting for temperature. Exploratory analysis did not reveal a statistically independent effect of humidity in the cities studied (Supplementary Section 2.1).

These observations motivate a more structured model that directly targets the epidemiologic quantity of interest: the temperature at which mortality risk begins to rise above its baseline. In the next subsection, we propose a structured threshold-based model that parameterizes the temperature–mortality association using temperature at lag 1, assuming the relationship is approximately constant up to an unknown threshold and increases thereafter.

### 1.3 Statistical Model

In temperature–mortality time-series analyses, daily death counts are typically modeled through overdispersed Poisson regression. A convenient way to accommodate overdispersion is through a negative binomial formulation. Under this specification, the observed mortality count *Y*_*t*_ at each observed time *t* = 1,…,*T* follows a negative binomial distribution,

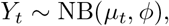

where *µ*_*t*_ = E(*Y*_*t*_) is the expected mortality count and *ω* is an overdispersion parameter,

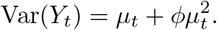

We used a log link to relate the expected death count *µ*_*t*_ to seasonal trends and temperature effects,

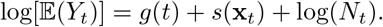

Here, *g*(*t*) is a smooth function capturing seasonal and long-term trends, *s*(**x**_*t*_) represents the temperature-mortality relationship with **x**_*t*_ = (*x*_*t*_, *x*_*t*→1_,…, *x*_*t→L*_) with *x*_*t*_ the temperature at time *t*, and *L* denoting the maximum lag at which temperature is assumed to affect mortality, and *N*_*t*_ is the at-risk population. In DLNMs, the temperature effect is modeled as a sum of lagged contributions:

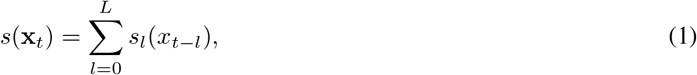

allowing temperatures on previous days to influence current mortality. Both *g*(*t*) and the *s*_*l*_(*x*_*t→l*_)s are specified using natural cubic splines (10). The function *g*(*t*) is assigned a fixed number of degrees of freedom per year to control for long-term and seasonal variation, while the *s*_*l*_(*x*_*t→l*_)s are defined using spline bases with knots placed at quantiles of the temperature distribution.

Motivated by the approximately flat temperature-mortality relationship at moderate temperatures, the sharp increase at high temperatures, and the strong correlation among lagged temperatures described in Section 1.2, we replaced the flexible representation used by DLNMs with a structured parameterization that directly targets the onset of heat-related excess mortality. Specifically, we assumed

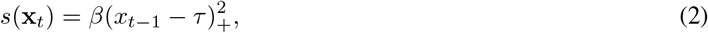

where (*u*)_+_ = max(*u,* 0), *ϑ* is an unknown threshold parameter representing the heat-risk onset temperature (HOT), and *ε* controls the rate at which mortality risk increases beyond this threshold. This specification uses temperature at lag 1, *x*_*t→1*_, motivated by exploratory data analysis indicating that a single lag captures most of the relevant variation in mortality risk. Under this model, the temperature-mortality association is constant for *x*_*t→1*_ ↓ *ϑ* and increases smoothly for *x*_*t→1*_ > *ϑ*, with larger *ε* values indicating greater sensitivity to heat above the threshold.

Note that the threshold parameter *ϑ* renders the model non-linear. To enable standard optimization algorithms, we approximate *s*(**x**_*t*_) with a smooth differentiable (Supplementary Section 2.2.1). Degrees of freedom for *g*(*t*) were similarly selected by visual inspection to capture seasonal patterns without absorbing short-term heat-related mortality signals (Supplementary Section 2.2.4). Given these model components, excess mortality for any set of days *A* is defined as 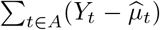 where 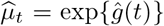 is the estimated baseline in the absence of heat effects. Days above the estimated HOT are further classified into low, moderate, and high risk categories based on *s*(**x**_*t*_), corresponding to increases in expected mortality of less than 5%, between 5% and 10%, and at least 10%, respectively (Supplementary Section 2.2.5. A simulation study based on Chennai data confirmed that the structured threshold model improves the bias–variance tradeoff relative to existing approaches (Supplementary Section 2.3).

## 2. Results

### 2.1 Heat-risk onset temperature estimates

We estimated the heat-risk onset temperature (HOT) for five Indian cities, along with the corresponding tempera-ture–mortality relationships, through a model specification that uses lag 1 temperature as predictor, *x*_*t→1*_ (Figure 2). This model choice was motivated by exploratory data analysis indicating that a single one day lagged temperature captured most of the relevant variation in mortality risk . Model fit was assessed using Pearson residual Q–Q plots (Supplementary Figure S21), which which were consisted with model assumptions.

**Figure 2.**
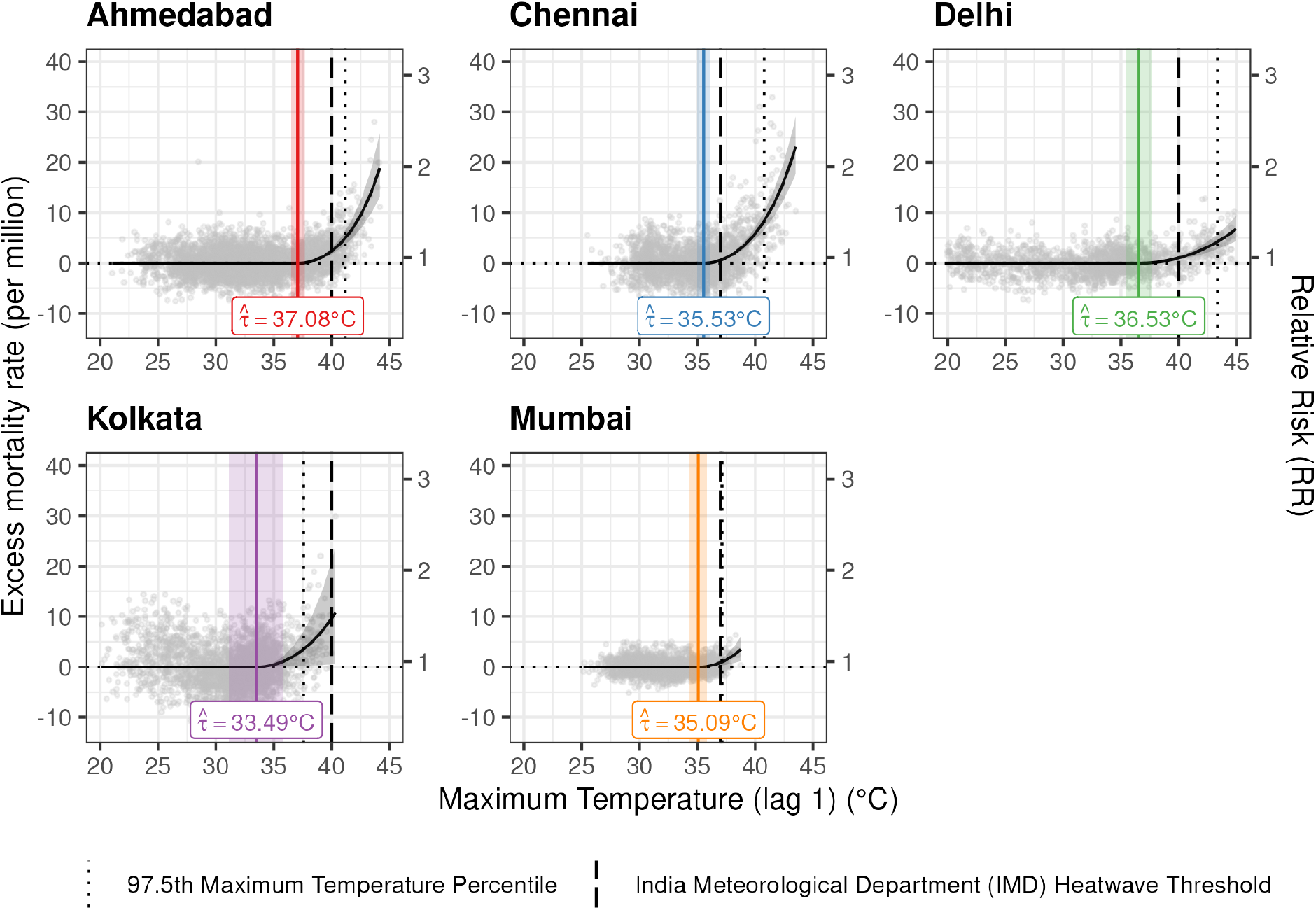
Estimated temperature thresholds for five Indian cities. Grey points represent residuals from a model fit that adjusted for long-term and seasonal patterns but excluded the temperature effect, represented as excess deaths per year per million, and the x-axis shows lag 1 maximum temperature. The vertical lines and corresponding shaded rectangle represent the estimated heat-risk onset temperature with 95% confidence intervals. The dashed-dot lines represent the maximum temperature threshold for heatwave currently used by the IMD and the dotted vertical lines represent the 97.5th percentile temperature of the observed maximum temperature distribution. For improved visualization, we limit the range of the x-axis to be 20-45, which excludes lower temperatures for Delhi.

Estimated HOT values were 37.1 °C (95% CI: 36.5–37.7) in Ahmedabad, 35.5 °C (35.0–36.1) in Chennai, 36.5 °C (35.4–37.7) in Delhi, 33.5 °C (31.2–35.8) in Kolkata, and 35.1 °C (34.3–35.8) in Mumbai. Estimated HOT values occurred within the upper range of the observed temperature distributions in each city.

Across cities, estimated HOT values were close to or below 37 °C and lower than the operational heat wave thresholds defined by the India Meteorological Department (IMD), which specify 37 °C for coastal cities (Chennai and Mumbai) and 40 °C for inland cities (Ahmedabad, Delhi, Kolkata).

At higher temperatures, mortality risk increased across cities. The magnitude of this increase varied, with the largest estimated effect in Ahmedabad 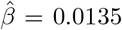, followed by Mumbai 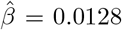, Chennai showed a moderate increase 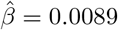, while smaller estimated effects were observed in Delhi 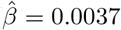 and Kolkata 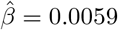

### 2.2 Excess deaths estimates

Our model finds substantial excess mortality associated with high temperatures. Based on the periods studied, we estimate annual excess deaths of 1,469, 1,369, 773, 580, and 411 for Chennai, Delhi, Ahmedabad, Kolkata, and Mumbai, respectively. The majority of excess deaths occurred during high-risk periods, with increases in excess mortality during these days (Table 1).

**Table 1:** Excess mortality by city and heat-risk category. Heat-risk categories (low, moderate, high) are defined based on estimated increases in mortality risk relative to baseline; an additional row per city reports results for a fixed IMD heatwave threshold. For each city and category, the table shows the corresponding temperature range, average number of heat-wave days per year, average number of days per year in runs of at least 3 and 7 consecutive days above the threshold, total excess deaths, and excess mortality rates per 100,000 persons per year (with 95% confidence intervals).

| City (Years of data) | Risk category<br>and temperature range | Days/yr | Days in<br>runs ≥3/yr | Days in<br>runs ≥7/yr | Total excess<br>deaths | Excess deaths per<br>100,000 per yr (95% CI) |
|---|---|---|---|---|---|---|
| **Ahmedabad (10)** | 0–5%: [37.08, 38.99) | 37.0 | 24.5 | 9.0 | 1,234 | 2.73 (2.64, 2.83) |
|  | 5–10%: [38.99, 39.75) | 13.0 | 4.1 | 0.0 | 787 | 1.77 (1.71, 1.82) |
|  | >10%: 39.75 + | 28.8 | 26.1 | 16.0 | 5,710 | 12.36 (12.28, 12.45) |
|  | Total: 37.08 + | 78.8 | 75.6 | 66.4 | 7,731 | 17.02 (16.88, 17.15) |
|  | IMD: 40.00 + | 25.1 | 22.3 | 13.9 | 5,311 | 11.49 (11.42, 11.57) |
| **Chennai (6)** | 0–5%: [35.53, 37.87) | 85.3 | 65.5 | 30.2 | 699 | 2.25 (2.01, 2.48) |
|  | 5–10%: [37.87, 38.80) | 23.5 | 6.3 | 1.2 | 1,612 | 5.16 (5.04, 5.29) |
|  | >10%: 38.80 + | 31.8 | 25.0 | 15.3 | 6,500 | 20.97 (20.83, 21.12) |
|  | Total: 35.53 + | 140.7 | 130.7 | 111.8 | 8,811 | 28.32 (28.02, 28.63) |
|  | IMD: 37.00 + | 83.7 | 74.0 | 55.7 | 8,594 | 27.63 (27.39, 27.86) |
| **Delhi (5)** | 0–5%: [36.53, 40.18) | 42.4 | 31.4 | 11.4 | 959 | 1.62 (1.50, 1.73) |
|  | 5–10%: [40.18, 41.63) | 10.0 | 1.2 | 0.0 | 649 | 1.10 (1.04, 1.15) |
|  | >10%: 41.63 + | 23.8 | 19.6 | 12.0 | 5,234 | 8.85 (8.77, 8.93) |
|  | Total: 36.53 + | 76.2 | 72.2 | 63.6 | 6,843 | 11.55 (11.40, 11.70) |
|  | IMD: 40.00 + | 35.8 | 32.8 | 26.0 | 5,969 | 10.08 (9.98, 10.18) |
| **Kolkata (8)** | 0–5%: [33.49, 36.38) | 90.2 | 71.2 | 34.4 | 2,245 | 5.55 (5.29, 5.80) |
|  | 5–10%: [36.38, 37.53) | 7.4 | 1.2 | 0.0 | 366 | 0.92 (0.84, 1.00) |
|  | >10%: 37.53 + | 9.5 | 7.9 | 3.5 | 2,026 | 5.08 (4.99, 5.16) |
|  | Total: 33.49 + | 107.1 | 92.6 | 64.0 | 4,637 | 11.48 (11.20, 11.76) |
|  | IMD: 40.00 + | 1.0 | 0.0 | 0.0 | 140 | 0.37 (0.36, 0.38) |
| **Mumbai (5)** | 0–5%: [35.09, 37.05) | 59.8 | 50.4 | 33.0 | 1,145 | 1.72 (1.61, 1.83) |
|  | 5–10%: [37.05, 37.83) | 9.0 | 4.8 | 3.2 | 608 | 0.89 (0.85, 0.94) |
|  | >10%: 37.83 + | 2.2 | 0.0 | 0.0 | 303 | 0.56 (0.54, 0.59) |
|  | Total: 35.09 + | 70.6 | 65.6 | 55.4 | 2,056 | 3.08 (2.96, 3.20) |
|  | IMD: 37.00 + | 12.2 | 7.6 | 4.0 | 958 | 1.41 (1.36, 1.46) |

In Ahmedabad, the excess mortality rate increased from 2.73 excess deaths per 100,000 persons per year (95% CI: 2.64–2.83) in the low-risk category to 12.36 (12.28–12.45) in the high-risk category. A similar pattern was observed in Chennai (2.25 to 20.97) and Delhi (1.62 to 8.85), indicating substantially higher rates during the hottest days. In Kolkata and Mumbai, low- and high-risk categories had comparable rates (Kolkata: 5.55 vs 5.08; Mumbai: 1.72 vs 0.56), likely because these cities experience fewer days in the highest risk range, especially Mumbai with only 2.2 high-risk days per year on average. In general, the increase in mortality rates was nonlinear, with the largest jump occurring between the moderate- and high-risk categories in most cities. Despite representing a smaller share of total days, high-risk periods accounted for the majority of excess deaths in three of the cities.

Differences in total excess deaths across cities are largely explained by differences in the number of high-risk days. Chennai, Ahmedabad, Delhi, Kolkata, and Mumbai experienced an average of 31.8, 28.8, 23.8, 9.5, and 2.2 high-risk days per year, respectively.

## 3. Discussion

We used a structured threshold model to estimate the temperature at which heat-related mortality risk rises above seasonal levels in five Indian cities. In all cities, the estimated heat-risk onset temperatures (HOTs) were lower than the operational heat wave thresholds used by the India Meteorological Department (IMD) (18). This implies that under current definitions, heat wave warnings based on maximum temperature are not issued until after mortality risk is already elevated. This is consistent with Indian and international evidence showing significant mortality risk accumulating well below IMD’s absolute temperature thresholds (21; 26; 27; 35).

Excess mortality increased with temperature in most cities. Higher-intensity heat days (defined as *>* 10% increased mortality risk above baseline), which are less common, accounted for a larger share of the excess mortality. This pattern appeared in all five cities, except Kolkata and Mumbai where extreme temperatures were less frequent compared to the others. Although overall excess mortality was lower in Kolkata and Mumbai, the relative increase in mortality at higher temperatures remained consistent across cities. Notably, the range of HOTs across cities is within 4 °C, raising the possibility of a common temperature threshold range at which heat stress begins to drive heat-related mortality in a homogeneous population. Disentangling climate, acclimatization, and vulnerability contributions requires study across diverse cities.

These findings make a case for revisiting India’s heatwave declaration thresholds, as these thresholds are significantly higher than the HOTs estimated. While current IMD thresholds may have operational advantages given the clear delineation of thresholds between inland, coastal, and hilly areas, there is a strong case for recalibrating these thresholds using epidemiological evidence. Heat warning systems designed around meteorological convention rather than observed mortality risk are likely to leave populations exposed and systematically under-protect the population during dangerous early-phase heat exposure. Emerging evidence shows that the microenvironments where those most at risk live, work, and sleep are hotter than recorded meteorological temperatures (15; 32; 24), a gap that may have widened in recent decades with growing urban density, heat island effects, and changes in construction practices.

Balancing epidemiological accuracy with operational manageability is a concern, however, with considerations of alert fatigue and state capacity to respond to alerts being of greatest relevance to policymakers (29). The tiered risk categorization of low, moderate, and high risk days above the HOT developed in this study offers one practical resolution. A graduated response ladder would allow government capacity to scale with risk level. Triggering public warning systems only at moderate or high risk levels would also ameliorate concerns of alert fatigue, as would, when feasible, calibrating warning thresholds to specific population risk groups.

The implications of these findings extend beyond national threshold-setting to the broader architecture of heat governance in India. The HOT estimates reported in this study provide the kind of city-specific evidence that can guide the development and implementation of tiered HAP responses contextualized for local conditions; a gap in heat policy in India currently (30). We note that the model used in our analysis did not include covariates to account for potential effect modifiers such as air pollution, age structure, or socioeconomic vulnerability. However the model can be readily adapted to incorporate these. The aggregated daily counts of death used in this study are all-cause totals, precluding disaggregation by age or sex. Systemic challenges with cause-of-death coding precludes disaggregation by specific causes.

Humidity was excluded from the final model, as exploratory analysis did not reveal a statistically independent effect on all-cause mortality in the cities studied (Supplementary Section 2.1), consistent with findings from multi-country analyses (11). This absence of an effect should not be interpreted as evidence that no association exists; the relatively short time series and all-cause mortality outcome likely limited statistical power to detect a humidity effect. Furthermore, the present analysis focuses on mortality and does not address the likely morbidity burden associated with combined heat and humidity exposure.

This study is part of a small but growing body of locally grounded heat epidemiology in India. The approach underlying this analysis, with mortality data obtained from municipalities, could feasibly be replicated across other cities in India. Extending this analytical framework across other cities, and strengthening the linkages between epidemiology and heat policy should be priorities for research and policy communities. Our results demonstrate that epidemiologically derived thresholds are feasible and informative across Indian cities. Aligning heat warning thresholds with observed mortality risk is a concrete, actionable step toward more effective heat-health warning systems, requiring collaboration between the epidemiological research community, the IMD, and disaster management authorities (1).

### Ethics approval

This study was reviewed by the Institutional Review Board (IRB) of the Harvard T.H. Chan School of Public Health and determined to be Not Human Subjects Research as defined by DHHS regulations (45 CFR 46.102(e)), under protocol number IRB24-1649. As this study uses only aggregate mortality data with no direct involvement of human subjects, additional ethics review was not required.

## Data Availability

The mortality data are not publicly available. Access may be possible through the relevant data custodians, subject to their approval. Daily maximum temperature data are available from the India Meteorological Department (gridded dataset), and absolute humidity data from NASA POWER. Code to fit the model is available in the HOTR R package at [https://github.com/roblesm15/hotR.git].

## Author contributions

MRF and RI conceptualized and designed the statistical methodology. MRF developed and implemented the methodology, conducted all analyses, and led the writing of the manuscript. RI supervised the study, provided critical structural guidance on the manuscript, and contributed substantially to the writing and revision of the manuscript. BK, AM, and AT provided the mortality data used in this study. BK and AM also cleaned, curated, validated the mortality data for all cities. All authors participated in drafting the manuscript or revising it critically for content. MRF is the guarantor of this work and takes responsibility for the integrity of the statistical methodology and the accuracy of the analysis.

## Supplementary data

Supplementary data are available online. Package implementation in R for the statistical model presented in this work is available here: HOTR.

## Use of artificial intelligence (AI) tools

AI tools (ChatGPT, OpenAI; Claude, Anthropic) were used during the preparation of this work to assist with writing clarity and code refinement. All AI-assisted content was reviewed and verified before integration.

