## Supplementary Data for "Data-driven estimation of the onset of heat-related mortality in five Indian cities"

### 2 Supplementary Sections

#### 2.1 Analyzing the effect of humidity

We obtained absolute humidity (AH) for each of the five Indian cities from the National Aeronautics and Space Administration (NASA) Langley Research Center's Prediction Of Worldwide Energy Resources (POWER) project, funded through the NASA Earth Science Division.

To isolate the effect of AH, we fit a model that accounts for long-term trends in mortality and compared the resulting residuals against both AH and maximum temperature. For all cities except Kolkata, the highest excess deaths per million population are observed near the center of the AH distribution rather than at its upper end. Only in Kolkata do we see some evidence of a joint temperature–humidity effect, with one data point lying in the upper portion of both distributions (Figure S1).

We further investigated humidity effects by evaluating whether any remaining variation in mortality could be explained by AH after accounting for seasonality, long-term trends, and maximum temperature. To isolate seasonal effects of humidity, we examined mortality residuals from the structured threshold model (using temperature at lag 1) and plotted them against AH. We overlaid a one-dimensional LOESS smoother to flexibly capture any trend. Across cities and months, the fitted curves are approximately flat, indicating little evidence of an independent association between humidity and mortality after accounting for temperature (Figure S2).

These results might suggest that temperature alone explains excess mortality associated with hot weather. However, the number of observed data points is small in each city, limiting our ability to draw firm conclusions about whether a humidity effect exists.

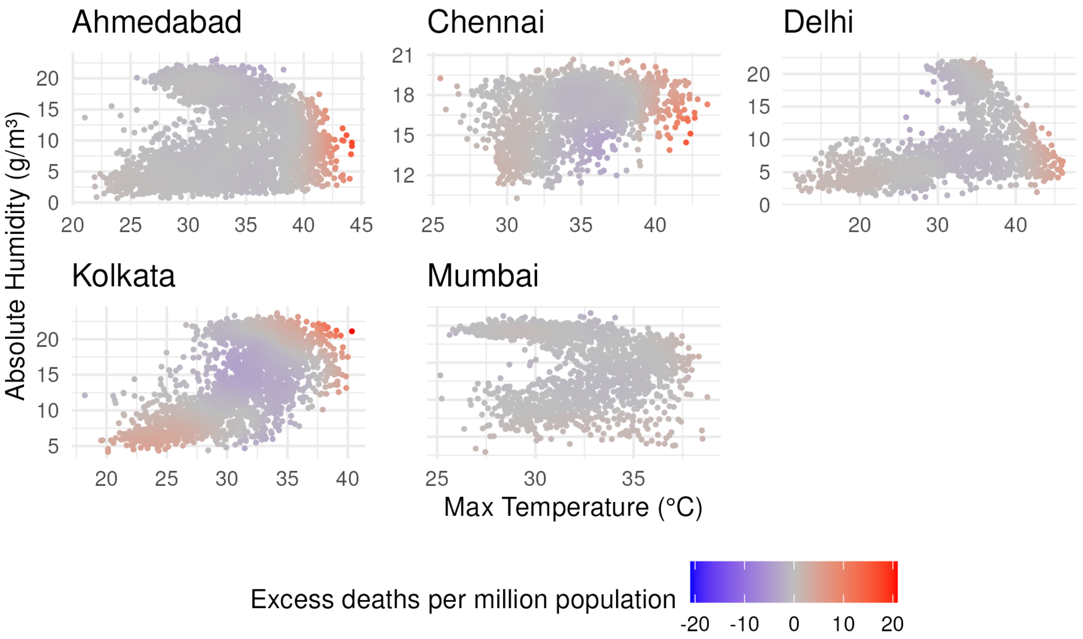


Supplementary Section Figure S1: Maximum temperature vs. absolute humidity colored by colored fitted LOESS estimate of residual mortality rate (per million). We model long-term and seasonal trends using natural cubic splines, with the number of degrees of freedom per year selected based on exploratory analysis (three for Ahmedabad, Chennai, Delhi, and Kolkata, and four for Mumbai). We extract the model residuals and use them to identify a joint effect of temperature and humidity, if any. We smooth the joint effect of temperature and humidity using a local quadratic polynomial (degree = 2) with span = 0.25 and use the fitted LOESS surface of residual mortality rate (per million) to color the points.

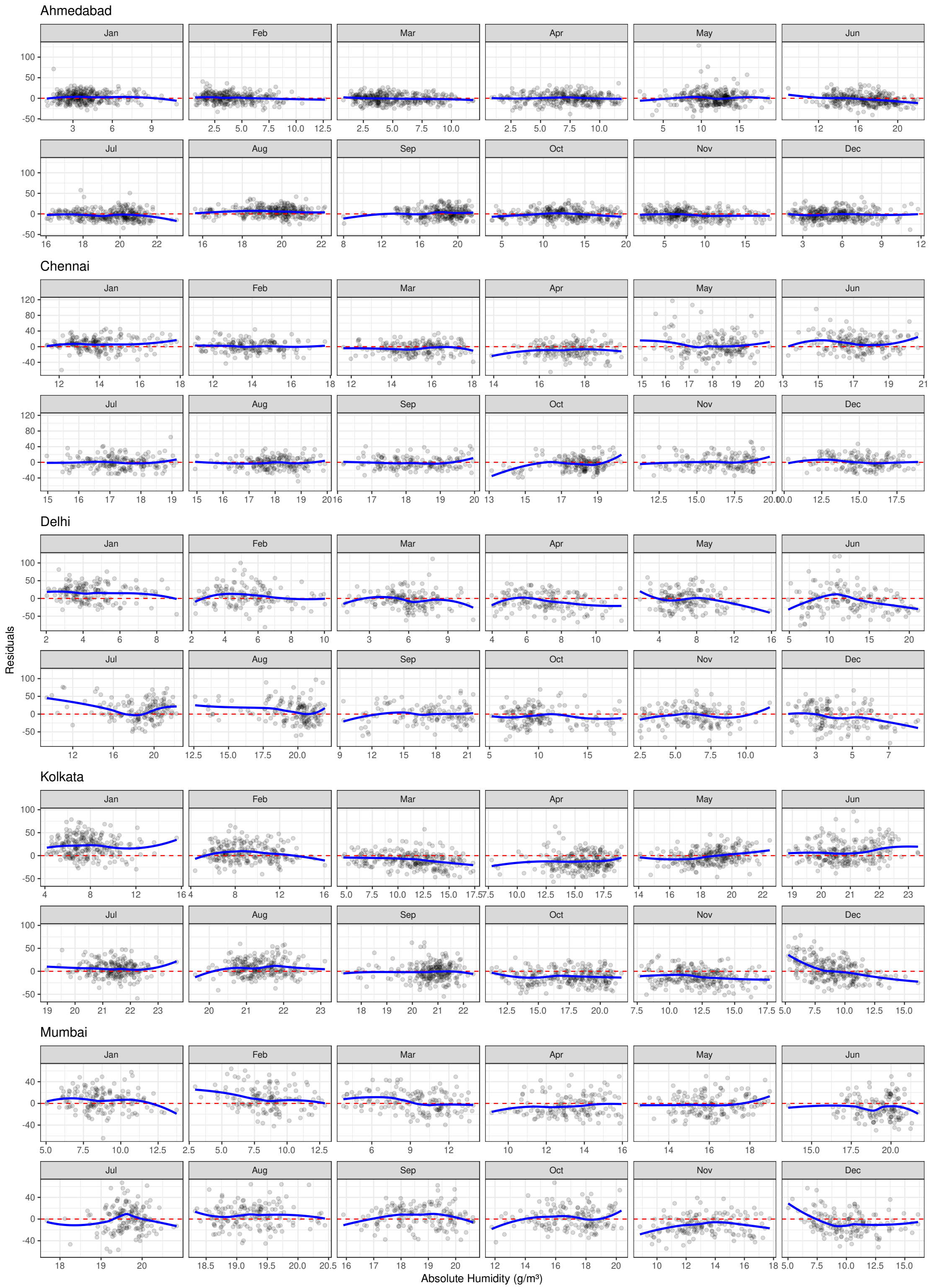


Supplementary Section Figure S2: Residual mortality from the structured threshold model (using temperature at lag 1) plotted against absolute humidity, by month, for each city. Absolute humidity data were obtained from the NASA POWER dataset. Monthly panels are shown to assess potential residual seasonality, as the parsimonious specification of the long-term trend may leave some seasonal structure to avoid overfitting heat-related variation. A LOESS smoother (default span, degree = 1) is overlaid in each panel.

#### 2.2 Methods details

##### 2.2.1 Estimation

Since smoothing splines can be expressed as linear combinations of known functions, the original model with (1) is a generalized linear model (GLM). Therefore, estimation can use standard maximum likelihood methods within the GLM framework.

However, we defined $s(\mathbf{x}_t) = \beta(x_{t-1} - \tau)_+^2$, a function not linear in the parameter $\tau$. Consequently, the model no longer falls within the GLM class, and maximum likelihood estimation must be carried out using general-purpose optimization algorithms. Another challenge is that $s(\mathbf{x}_t)$ is not smooth at $x_t = \tau$, limiting the use of standard second-order optimization methods. To address these, we approximate $s(\mathbf{x}_t)$ with a smooth and differentiable function that permitted standard optimization algorithms be applied.

##### 2.2.2 Smooth approximation

$s(x_t; \beta)$ is not smooth at $x_t = \tau$. As a result, derivatives with respect to $\tau$ are not well-behaved, and the log-likelihood is not locally quadratic. This limits the use of standard second-order optimization methods, such as Newton-Raphson.

To address this challenge, we approximated $s(x_t; \beta)$ with the following smooth and differentiable function:

$$s(\mathbf{x}_t) \approx \beta(x_{t-1} - \tau)^2 \cdot \frac{1}{1 + e^{-k(x_{t-1}-\tau)}} \tag{3}$$

Here, $k > 0$ is a parameter that adjusts how quickly the transition happens near $\tau$. As $k$ increases, the approximation gets closer to $\beta(x_{t-1} - \tau)_+^2$. In practice, $k = 5$ yields results that are almost identical to those of the original function specification (2).

With this smooth approximation in place, we applied standard gradient-based optimization algorithms and computed the necessary derivatives for estimation and inference, including standard errors.

##### 2.2.3 Optimization algorithm

We estimate the parameter vector

$$\theta = (\boldsymbol{\gamma}, \beta, \tau, \phi)$$

by maximizing the log-likelihood $\ell(\theta)$ using a hybrid block coordinate algorithm.

**Algorithm:**

1. **Initialization.**
   - Initialize the threshold $\tau^{(0)}$.
   - Construct the design matrix for the intercept and time spline.
   - Fit an initial negative binomial model using `glm.nb` to obtain starting values for $(\boldsymbol{\gamma}, \beta, \phi)$.
2. **Iterative updates.** For $i = 0, 1, 2, \ldots$ until convergence:
   (a) Given $\tau^{(i)}$, update $(\boldsymbol{\gamma}, \beta)$ using a negative binomial GLM with dispersion held fixed.
   (b) Update $\tau$ by maximizing the log-likelihood with the other parameters fixed.
   (c) Periodically refresh the dispersion parameter $\phi$ while holding the other parameters fixed.
3. **Final update.** After convergence, perform one final update of the dispersion parameter.
4. **Convergence criterion.** Stop when the change in the log-likelihood or parameters falls below a predefined tolerance.

##### 2.2.4 Choice of degrees of freedom per year

When splines are used to explain long-term trends, selecting the degrees of freedom for $g(t)$ is critical, as overly flexible specifications can absorb short-term increases in mortality associated with heat events, attenuating the estimated temperature effect. We selected the degrees of freedom based on visual inspection of model fits, aiming for sufficient flexibility to capture seasonal patterns while avoiding responsiveness to short-term fluctuations driven by heat episodes (Supplementary Figure S20). We used three degrees of freedom per year for Ahmedabad, Chennai, Delhi, and Kolkata, and four degrees of freedom for Mumbai.

##### 2.2.5 Excess mortality

For any subset of days $A$, we define observed excess mortality as the difference between observed mortality and baseline expected mortality:

$$\sum_{t \in A} (Y_t - \mu_t),$$

where $Y_t$ is the observed number of deaths on day $t$ and $\mu_t = \exp\{g(t)\}$ represents the expected (baseline) mortality in the absence of heat effects. In practice, we estimate this quantity using fitted model components:

$$\sum_{t \in A} (Y_t - \widehat{\mu}_t),$$

where $\widehat{\mu}_t = \exp\{\hat{g}(t)\}$ denotes the estimated baseline mortality. Our model-based approach also allows us to compute standard errors and confidence intervals for these quantities. To enable comparisons across cities with different population sizes and exposure durations, we can express excess mortality as a rate per person-time:

$$\frac{\sum_{t \in A} (Y_t - \widehat{\mu}_t)}{\sum_{t \in A} N_t},$$

where $N_t$ is the population at risk on day $t$.

A natural choice of subset for evaluating high temperature–related mortality is the set of days for which temperature exceeds the estimated HOT, i.e., $A = \{t : x_{t-1} \geq \hat{\tau}\}$. However, defining alerts based solely on this threshold may be impractical, for example due to warning fatigue, so summaries of interest may instead consider excess mortality across different levels of predicted risk. For example, we divide days above the estimated HOT into low, moderate, and high risk categories using the estimated temperature-mortality relationship.

Specifically, because in our approach $s(\mathbf{x}_t)$ represents the log-relative risk associated with temperature at lag 1 relative to baseline, it can be used to define risk-based categories. For each day $t$, we classify it into one of three levels of elevated risk:

$$0 < s(\mathbf{x}_t) < \log(1.05), \quad \log(1.05) \leq s(\mathbf{x}_t) < \log(1.10), \quad s(\mathbf{x}_t) \geq \log(1.10),$$

corresponding to increases in expected mortality of less than 5%, between 5% and 10%, and at least 10%, respectively. We refer to these categories as low, moderate, and high risk.

#### 2.3 Simulation

##### 2.3.1 Design

To evaluate whether the proposed approach improves upon existing methods, we conducted a simulation study. Because no dataset with known ground truth is available, simulation is required to assess estimation accuracy. To avoid bias in favor of the proposed model, we generated data under multiple data-generating processes, including two distributed lag nonlinear models (DLNMs) and the proposed structured threshold model.

Specifically, we considered three data-generating processes: (i) a DLNM in which temperature at the previous 2 days affects mortality, (ii) a DLNM in which temperature over the previous 7 days affects mortality, and (iii) our proposed structured threshold model. For (iii) we considered one value $\beta$. This design allows evaluation of performance both when the proposed model is correctly specified and when the true data-generating mechanism follows a more flexible DLNM structure.

To mimic real data, all simulation components were derived from observed data in Chennai, which was selected as representative of the temperature range and temporal structure in our study. We used the observed temperature series directly in the simulations. The long-term and seasonal mortality trend was estimated from the Chennai data and used as the underlying temporal component in all data-generating processes.

For the DLNM-based data-generating processes, we fit models to the Chennai data using natural cubic splines with 3 degrees of freedom per year for the seasonal trend and a cross-basis specification for temperature. We constructed two versions of the temperature–mortality relationship: one with 2 lagged days and one with 7 lagged days contributing to mortality. In both cases, the estimated spline coefficients from these fitted models were treated as the true parameters in the simulation (see Supplementary data sections 2.3.2 and 2.3.3).

For the threshold-based data-generating process, we generated data using the proposed model with $\tau = 35.5$°C and $\beta = 0.01$, using temperature at lag 1 ($x_{t-1}$) as the exposure. These values are broadly similar to the estimated parameters for Chennai. The same estimated seasonal trend was used as in the DLNM-based simulations.

Given the specified mean structure for each data-generating process, mortality counts were generated from a negative binomial distribution, with the overdispersion parameter set equal to the estimate obtained from the corresponding fitted model.

We generated 2,000 simulated datasets under each data-generating process.

##### 2.3.2 Long term trend: spline parameter specification

We used three degrees of freedom per year of data in Chennai, for a total of 18 spline basis column coefficients $\gamma = (-0.168,\ -0.054,\ -0.191,\ -0.198,\ -0.021,\ -0.315, -0.238,\ 0.012,\ -0.272,\ -0.291,\ -0.058,\ -0.235, -0.329,\ -0.056,\ -0.292,\ -0.053,\ -0.339,\ 0.046)$.

##### 2.3.3 Flexible model temperature–mortality curve spline parameter specification

We used 4 equally spaced knots placed within temperature range, for each lag. For a specification with 2 lags we used the following coefficients for the spline basis

$\boldsymbol{\beta} = (-0.110,\ -0.094,\ -0.070,\ -0.014,\ -0.013,\ -0.011, -0.027,\ -0.018,\ 0.008,\ 0.161,\ 0.071,\ 0.046)$.

Similarly, for the model with 7 lags, we used the following coefficients,

$\boldsymbol{\beta} = (-0.062,\ -0.053,\ -0.032,\ -0.041,\ -0.015,\ -0.026, -0.017,\ -0.010,\ -0.009,\ -0.011,\ -0.016,\ -0.003,\ -0.010,\ 0.004,\ -0.022,\ 0.037,\ 0.031,\ 0.025, 0.020,\ 0.103,\ 0.067,\ 0.094,\ 0.015,\ 0.147,\ 0.163,\ 0.063,\ 0.010,\ 0.188,\ 0.122,\ 0.140, 0.159,\ 0.199)$.

##### 2.3.4 Intercept adjustment

The DLNM does not constrain the temperature–mortality function to be centered at zero; baseline mortality is absorbed into the model intercept. Consequently, the estimated function $\hat{s}(\mathbf{x}_t)$ may be shifted vertically relative to the true function $s(\mathbf{x}_t)$ by a constant equal to the difference between the estimated and true intercepts. Before computing bias, we therefore adjust each replicate's estimated function by subtracting this intercept difference:

$$\hat{s}^*(\mathbf{x}_t) = \hat{s}(\mathbf{x}_t) - (\hat{\alpha} - \alpha),$$

where $\hat{\alpha}$ and $\alpha$ are the estimated and true intercepts, respectively.

##### 2.3.5 Model assessment

Model performance was assessed based on mean squared error (MSE) which we divided into three components:

$$\frac{1}{T}\sum_{t=1}^{T}\underbrace{\mathbb{E}\left[(\hat{s}(\mathbf{x}_t) - s(\mathbf{x}_t))^2\right]}_{\text{MSE}} = \{\underbrace{\mu_{\hat{s}} - \mu_s}_{\text{overall bias}}\}^2 + \frac{1}{T}\sum_{t=1}^{T}\left\{\underbrace{\mathbb{E}[\hat{s}(\mathbf{x}_t) - \mu_{\hat{s}}] - [s(\mathbf{x}_t) - \mu_s]}_{\text{shape bias}}\right\}^2 + \frac{1}{T}\sum_{t=1}^{T}\underbrace{\text{Var}\left[\hat{s}(\mathbf{x}_t)\right]}_{\text{variance}}$$

with the mean levels defined by:

$$\mu_s = \frac{1}{T}\sum_{t=1}^{T} s(\mathbf{x}_t) \text{ and } \mu_{\hat{s}} = \frac{1}{T}\sum_{t=1}^{T}\mathbb{E}[\hat{s}(\mathbf{x}_t)].$$

The expectations and variances are taken with respect to the distribution induced by the simulation replicates. Prior to computing MSE, estimated functions were adjusted for differences between the estimated and true model intercepts; details are provided in the Supplementary Data section 2.3.4.

The overall bias is not practically relevant because the temperature–mortality relationship is interpreted in relative terms, so constant shifts do not affect inference. In contrast, shape bias has important practical consequences, as it can alter key features of the curve, such as the location of the minimum of $s(\mathbf{x}_t)$. Variance is also critical, particularly for DLNMs, where higher variance leads to increased local fluctuations in the estimated curve, which can be misinterpreted as meaningful features, including spurious minima.

##### 2.3.6 Simulation results

For each simulated dataset, we fit DLNM specifications commonly used in practice ((31; 17; 20; 34)), with the temperature-mortality relationship defined by (1). Following standard practice, we considered models with different numbers of lags $L$, specifically 2, 7, 10, and 14 days, and used 3 degrees of freedom per year for the seasonal component in all cases. We also fit the proposed structured threshold model with the temperature-mortality relationship defined by (2).

Supplementary Section Table S1: Shape bias$^2$ and variance of the estimated temperature–mortality function over the simulation period. Shape bias$^2$ denotes the squared deviation of the estimated temperature–mortality function from the true function after centering, and variance denotes the variability of the estimated function across simulation replicates. Results are based on 2,000 simulated datasets per data-generating process. Columns correspond to the data-generating process: DLNM columns simulate a temperature–mortality effect via lag-2 or lag-7 distributed lag structures; the structured threshold model column uses a threshold effect with $\beta = 0.01$ at fixed $\tau = 35.5°\text{C}$. Row labels identify the fitting model: lag window for DLNM fits, or the structured threshold model fit using temperature at the previous day $x_{t-1}$.

| | | | DLNM | | Structured threshold model |
|---|---|---|---|---|---|
| | Model | Lag Structure | Lag 2 | Lag 7 | $\beta = 0.01$ |
| *Shape bias*$^2$ | DLNM | $\{x_{t-l}\}_{l=0}^{2}$ | 0.0000001 | 0.0009377 | 0.0000044 |
| | | $\{x_{t-l}\}_{l=0}^{7}$ | 0.0000002 | 0.0000006 | 0.0000045 |
| | | $\{x_{t-l}\}_{l=0}^{10}$ | 0.0000002 | 0.0000006 | 0.0000047 |
| | | $\{x_{t-l}\}_{l=0}^{14}$ | 0.0000003 | 0.0000010 | 0.0000047 |
| | Structured threshold model | $x_{t-1}$ | 0.0014242 | 0.0033311 | $\approx 0$ |
| *Variance* | DLNM | $\{x_{t-l}\}_{l=0}^{2}$ | 0.0024493 | 0.0026260 | 0.0024498 |
| | | $\{x_{t-l}\}_{l=0}^{7}$ | 0.0050470 | 0.0049578 | 0.0046033 |
| | | $\{x_{t-l}\}_{l=0}^{10}$ | 0.0064604 | 0.0062266 | 0.0057301 |
| | | $\{x_{t-l}\}_{l=0}^{14}$ | 0.0080456 | 0.0082552 | 0.0077244 |
| | Structured threshold model | $x_{t-1}$ | 0.0000215 | 0.0000245 | 0.0000275 |

Across all simulation settings, the structured threshold model produces substantially more stable estimates of the temperature–mortality relationship than DLNMs. The variability of the estimated heat function, even when the model is misspecified (Table S1) is substantially lower. In contrast, DLNMs exhibit substantially higher variability, which increases with model flexibility and the number of lags. As a result, the estimated curves from DLNMs are less stable across replicates, whereas the structured threshold model consistently recovers a smooth and interpretable relationship.

When the structured threshold model is misspecified (i.e., when the data-generating process follows a DLNM), it exhibits some shape bias in the estimated heat function (Table S1). This bias is expected, as the model constrains the temperature effect to be non-negative above the threshold, whereas DLNMs allow more flexible shapes. However, despite this constraint, the structured threshold model reliably recapitulates the overall shape of the temperature–mortality relationship. When the model matches the data-generating process, it provides unbiased and substantially more precise estimates of the parameter of interest, namely the heat-risk onset temperature $\tau$, compared to DLNMs estimating the minimum mortality temperature (MMT) (Table S2). Note that the mean CI width for DLNM MMT estimates is inflated by replicates with wide bootstrap confidence intervals. Overall, these results indicate that the structured threshold model improves the bias–variance tradeoff of current approaches, resulting in stable and interpretable estimates across a range of data-generating scenarios.

Supplementary Section Table S2: Standard deviation (SD), 95% confidence interval (CI) coverage, and mean CI width of the estimated parameter of interest across simulation replicates. Results are based on 2,000 simulated datasets per data-generating process. For DLNM fits the parameter of interest is the MMT; for the structured threshold model it is $\hat{\tau}$ at $\beta = 0.01$, $\tau = 35.5\,°\mathrm{C}$. Confidence intervals are percentile bootstrap-based for DLNMs and Wald-based for the structured threshold model. Dashes indicate model–data-generating process combinations that were not evaluated. Column and row definitions follow Table S1. Note that the mean CI width for DLNM MMT estimates is inflated by replicates with wide bootstrap confidence intervals.

| | | DLNM | | | | | | Structured threshold model | | |
|---|---|---|---|---|---|---|---|---|---|---|
| | | Lag 2 | | | Lag 7 | | | $\beta = 0.01$ | | |
| Model | Lag Structure | SD | Cov. | Width | SD | Cov. | Width | SD | Cov. | Width |
| DLNM | $\{x_{t-l}\}_{l=0}^{2}$ | 0.258 | 0.95 | 1.91 | 0.742 | 0.10 | 3.09 | | — | |
| | $\{x_{t-l}\}_{l=0}^{7}$ | 0.928 | 0.94 | 4.43 | 0.318 | 0.94 | 1.61 | | — | |
| | $\{x_{t-l}\}_{l=0}^{10}$ | 1.293 | 0.94 | 5.27 | 0.355 | 0.94 | 2.11 | | — | |
| | $\{x_{t-l}\}_{l=0}^{14}$ | 1.642 | 0.94 | 6.20 | 0.689 | 0.95 | 2.95 | | — | |
| Structured threshold model | $x_{t-1}$ | | — | | | — | | 0.301 | 0.94 | 1.12 |

### 2 Supplementary table

Supplementary Table S1: Summary of non-temperature-related high-mortality events in five Indian cities. Events were identified through systematic review of news reports and include natural disasters, infectious disease outbreaks, and other acute incidents associated with elevated mortality. For each event, the table reports the city, event date period, length (in days), and event type.

| City | Period | Length (days) | Event Type |
|---|---|---|---|
| Ahmedabad | 2001-01-26 to 2001-01-26 | 1 | Earthquake |
| Ahmedabad | 2002-02-28 to 2002-02-28 | 1 | Mob violence |
| Ahmedabad | 2002-09-24 to 2002-09-24 | 1 | Terrorist attack |
| Ahmedabad | 2006-08-15 to 2006-10-06 | 46 | Chikungunya |
| Ahmedabad | 2008-01-28 to 2008-02-11 | 15 | Jaundice |
| Ahmedabad | 2008-07-26 to 2008-07-26 | 1 | Bomb blast |
| Chennai | 2015-11-01 to 2015-12-31 | 61 | Flooding |
| Chennai | 2017-09-05 to 2017-10-05 | 31 | Dengue |
| Delhi | 2015-08-29 to 2015-09-28 | 31 | Dengue |
| Delhi | 2016-09-10 to 2016-10-10 | 31 | Chikungunya |
| Delhi | 2010-11-15 to 2010-11-15 | 1 | Building Collapse |
| Kolkata | 2010-03-23 to 2010-03-23 | 1 | Building Fire |
| Kolkata | 2011-12-09 to 2011-12-09 | 1 | Hospital fire |
| Kolkata | 2013-12-31 to 2014-01-15 | 16 | Dengue Fever |
| Kolkata | 2017-07-15 to 2017-08-14 | 31 | Floods |
| Kolkata | 2020-07-05 to 2020-08-14 | 41 | COVID-19 |
| Mumbai | 2010-07-15 to 2010-09-13 | 61 | Monsoon illness, Swine Flu, Malaria |

### 3 Supplementary figures

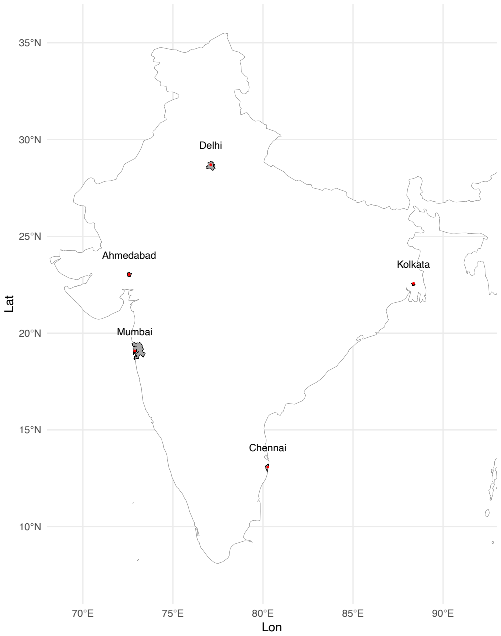


Supplementary Figure S1: Grid coordinates selected for each city's weather data. We used daily maximum temperature data from the India Meteorological Department's 1° latitude × 1° longitude gridded temperature dataset covering the period 1951–2018. For each city, we selected the grid cell whose centroid was closest to the urban core and that provided continuous coverage over the study period.

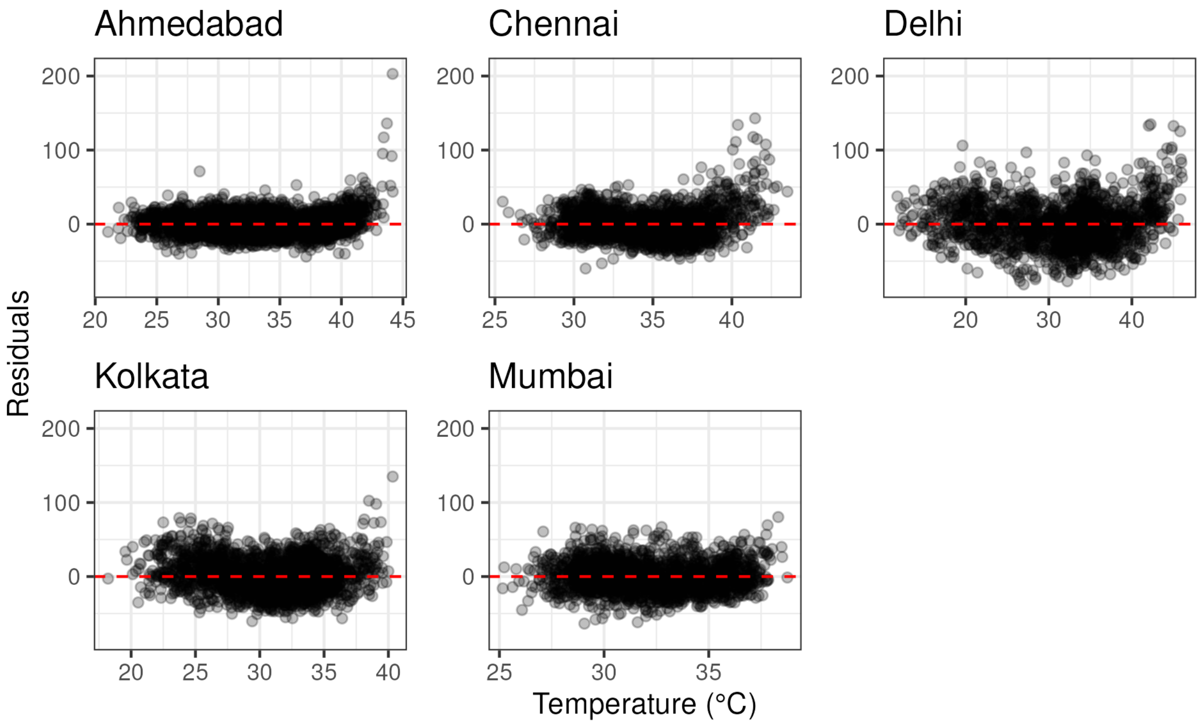


Supplementary Figure S2: Simple model residuals vs. temperature for five Indian cities. We model long-term and seasonal trends using natural cubic splines, with the number of degrees of freedom per year selected based on exploratory analysis (three for Ahmedabad, Chennai, Delhi, and Kolkata, and four for Mumbai). We extract the model residuals and plot them against temperature. Across cities, residuals show a systematic pattern: increase sharply at higher temperatures. This curvature and apparent "hinge" at high temperatures motivates the use of a nonlinear temperature function.

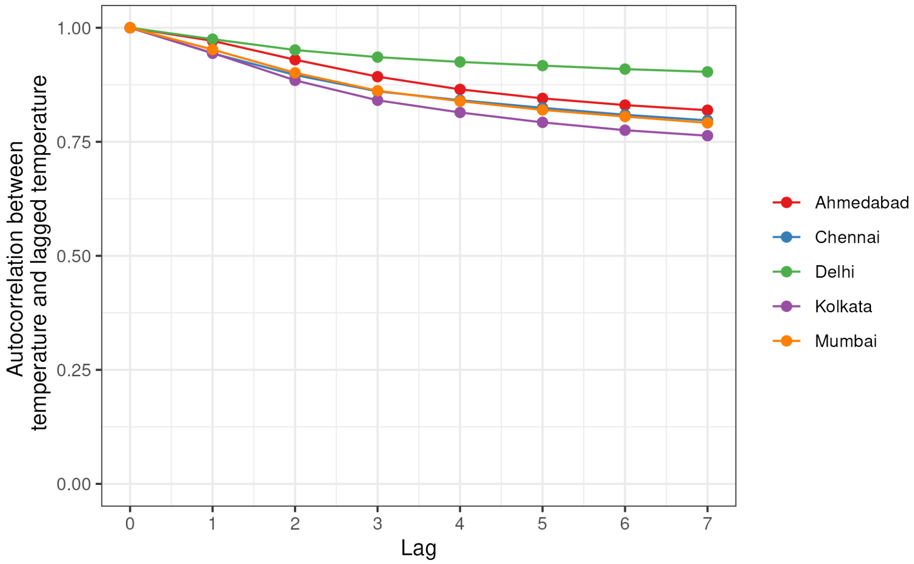


Supplementary Figure S3: Empirical autocorrelation for lagged temperatures. For all cities we compute the autocorrelation for each lag and. In all cities, correlations between lagged temperatures exceed 0.80, indicating strong dependence across lags.

**Ahmedabad**

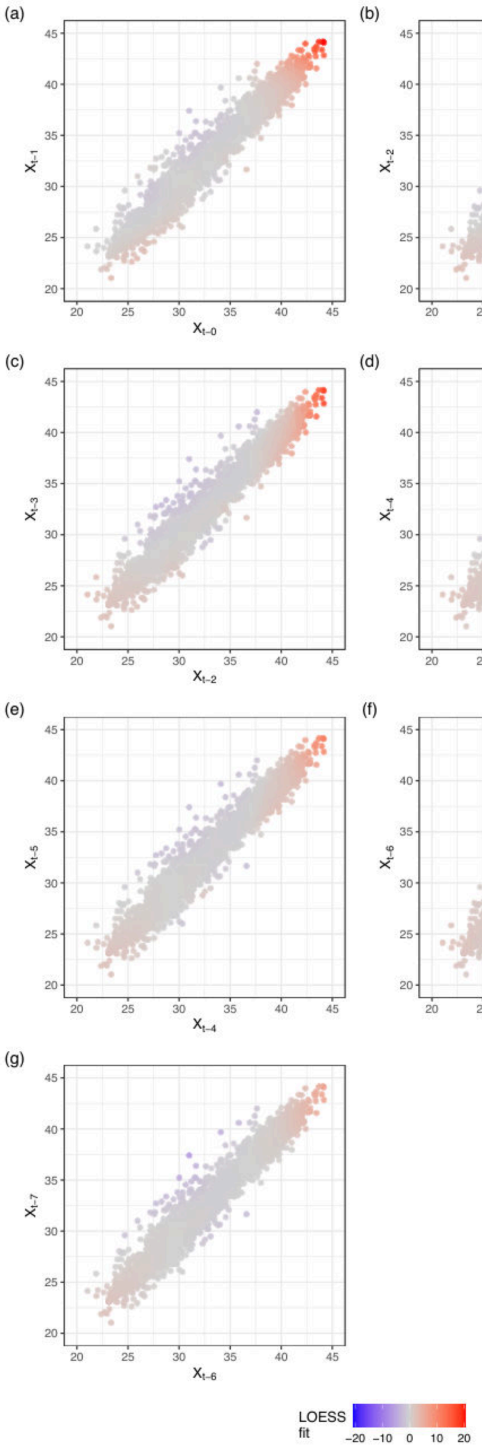


Supplementary Figure S4: Ahmedabad: Adjacent lagged temperatures colored fitted LOESS estimate of residual mortality rate. For each pair of adjacent temperature lags, fit a two-dimensional LOESS smoother to model residual mortality rate (from a model that accounts for the long-term and seasonal trend in mortality) as a function of the temperatures at times $t - l$ and $t - (l + 1)$ for $l \in \{0, 1, 2, 3, 4, 5, 6\}$. The smoother uses a quadratic local polynomial (degree = 2) with span = 0.25. Panels (a)–(g) compare adjacent lag pairs, with point color given by the fitted LOESS surface of residual mortality rate (per million). The heat effect is apparent in the upper right and mostly dissipated within 2 lags. Remaining cold day effects are due to parsimonious fit of long-term trend.

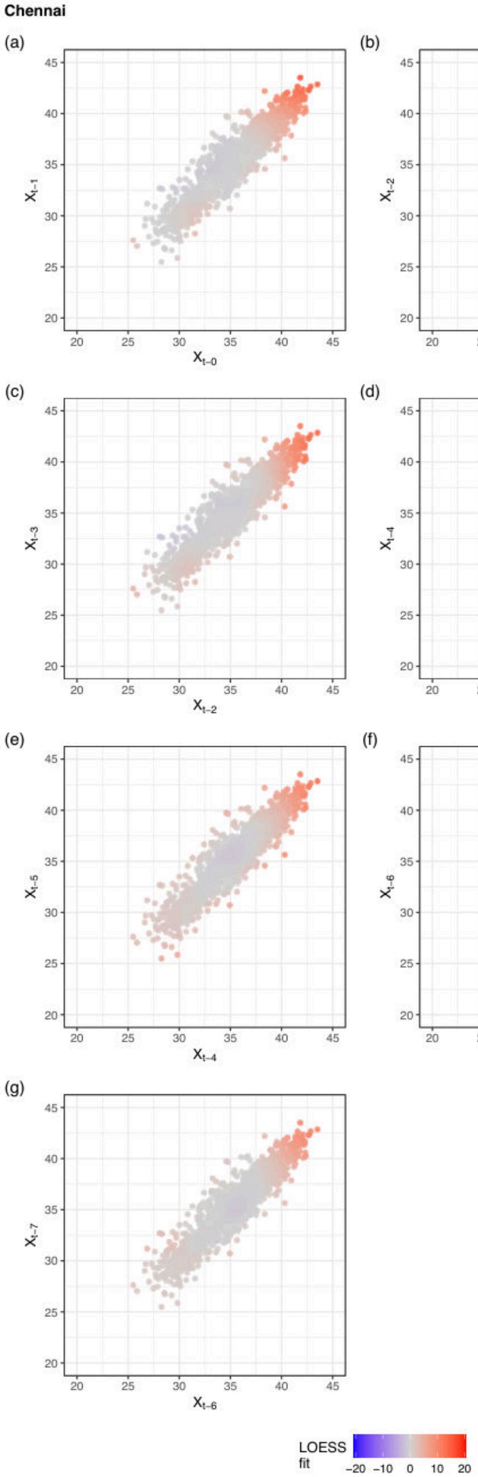


Supplementary Figure S5: Chennai: Adjacent lagged temperatures colored fitted LOESS estimate of residual mortality rate. See Figure S4 for full caption.

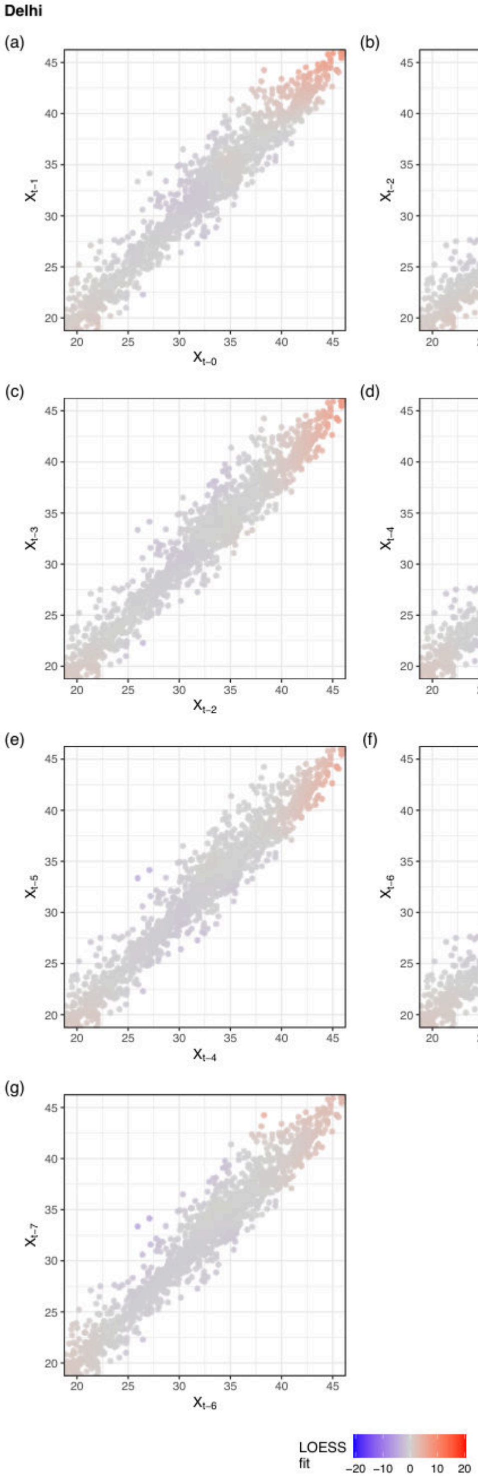


Supplementary Figure S6: Delhi: Adjacent lagged temperatures colored fitted LOESS estimate of residual mortality rate. See Figure S4 for full caption.

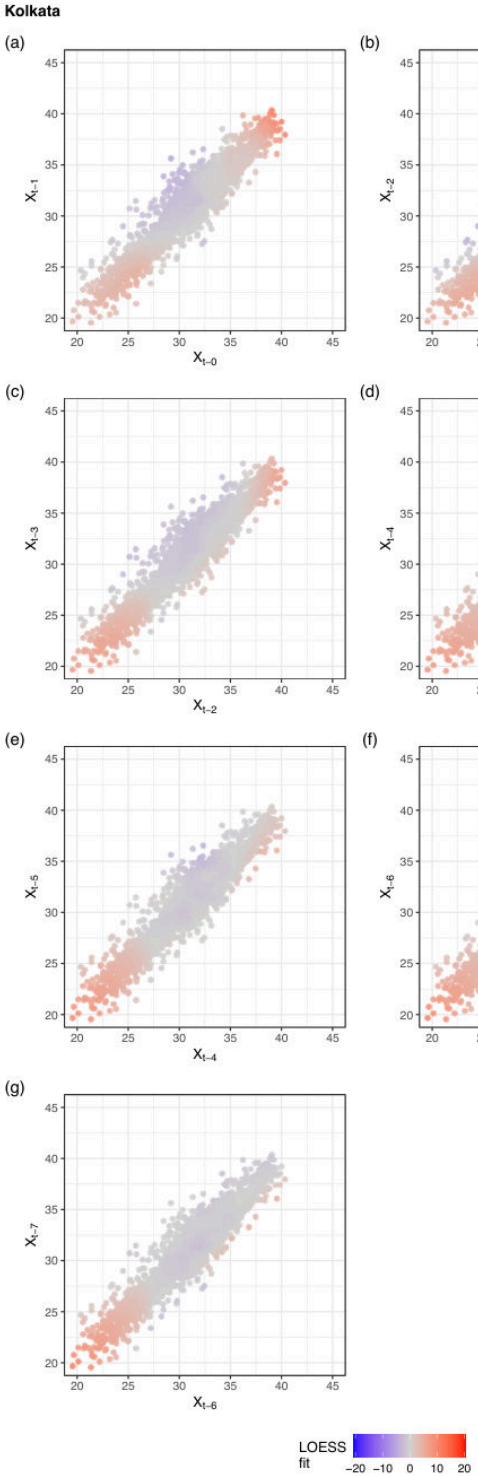


Supplementary Figure S7: Kolkata: Adjacent lagged temperatures colored fitted LOESS estimate of residual mortality rate. See Figure S4 for full caption.

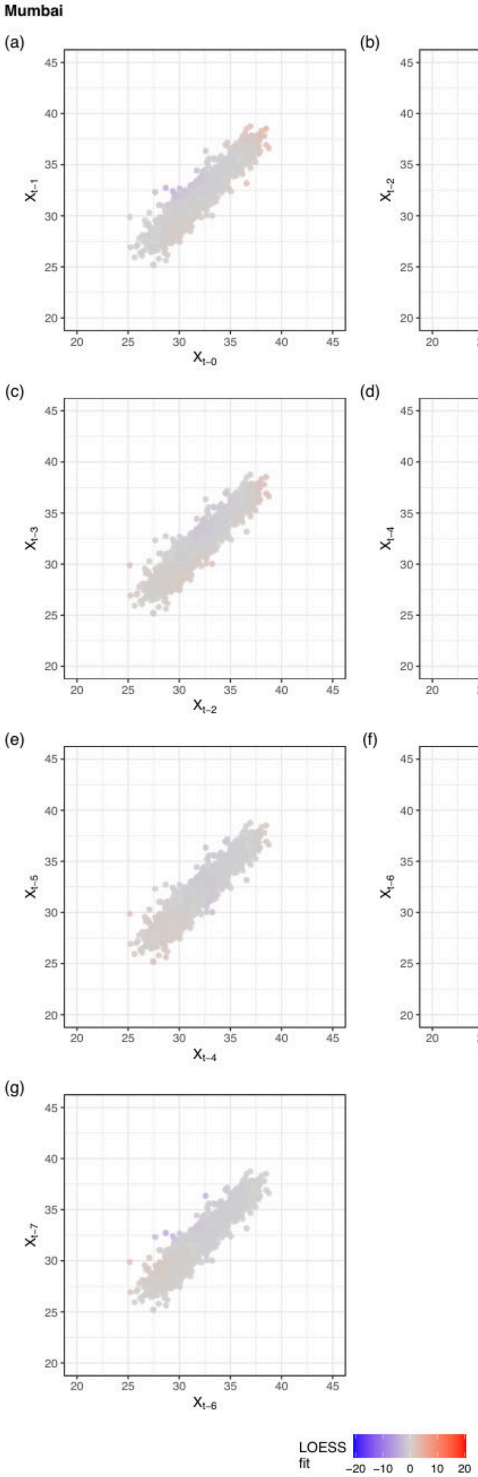


Supplementary Figure S8: Mumbai: Adjacent lagged temperatures colored fitted LOESS estimate of residual mortality rate. See Figure S4 for full caption.

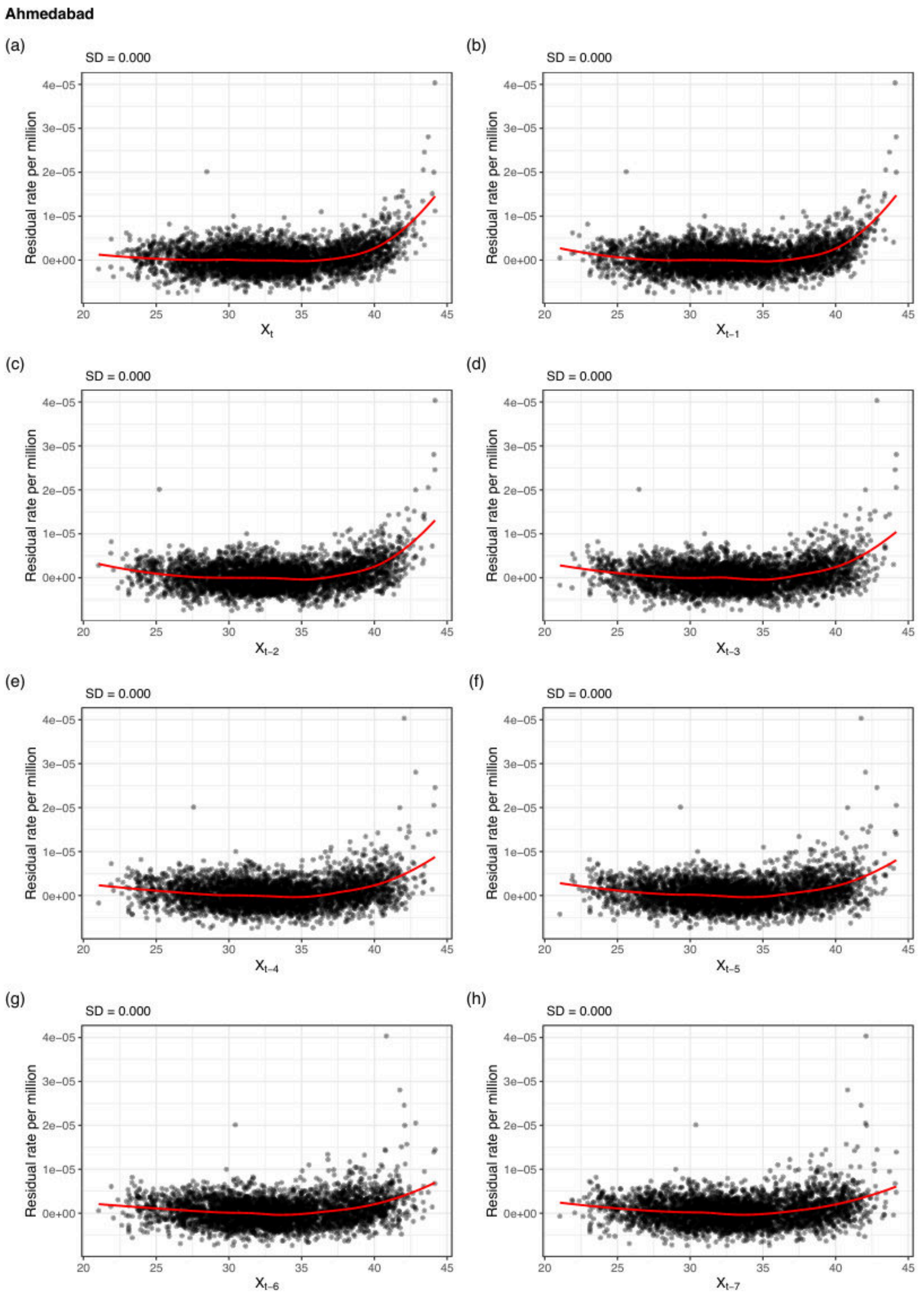


Supplementary Figure S9: Ahmedabad: Long-term fit residuals rate per million versus temperature at each lag. For each lag, we plot residual mortality rates (per million person-days), obtained from a model that accounts for long-term and seasonal trends in mortality, against temperature at time $t - l$, for $l \in \{0, 1, 2, 3, 4, 5, 6, 7\}$. For each lag, we fit a univariate LOESS smoother to model residual mortality rate as a function of temperature, using a local linear fit (degree = 1) with span = 0.5. Panels (a)–(h) show lag-specific residuals and corresponding LOESS smooths. We summarize the variability of each fitted curve using its standard deviation and find that lag 1 consistently explains the largest share of variation in mortality across cities.

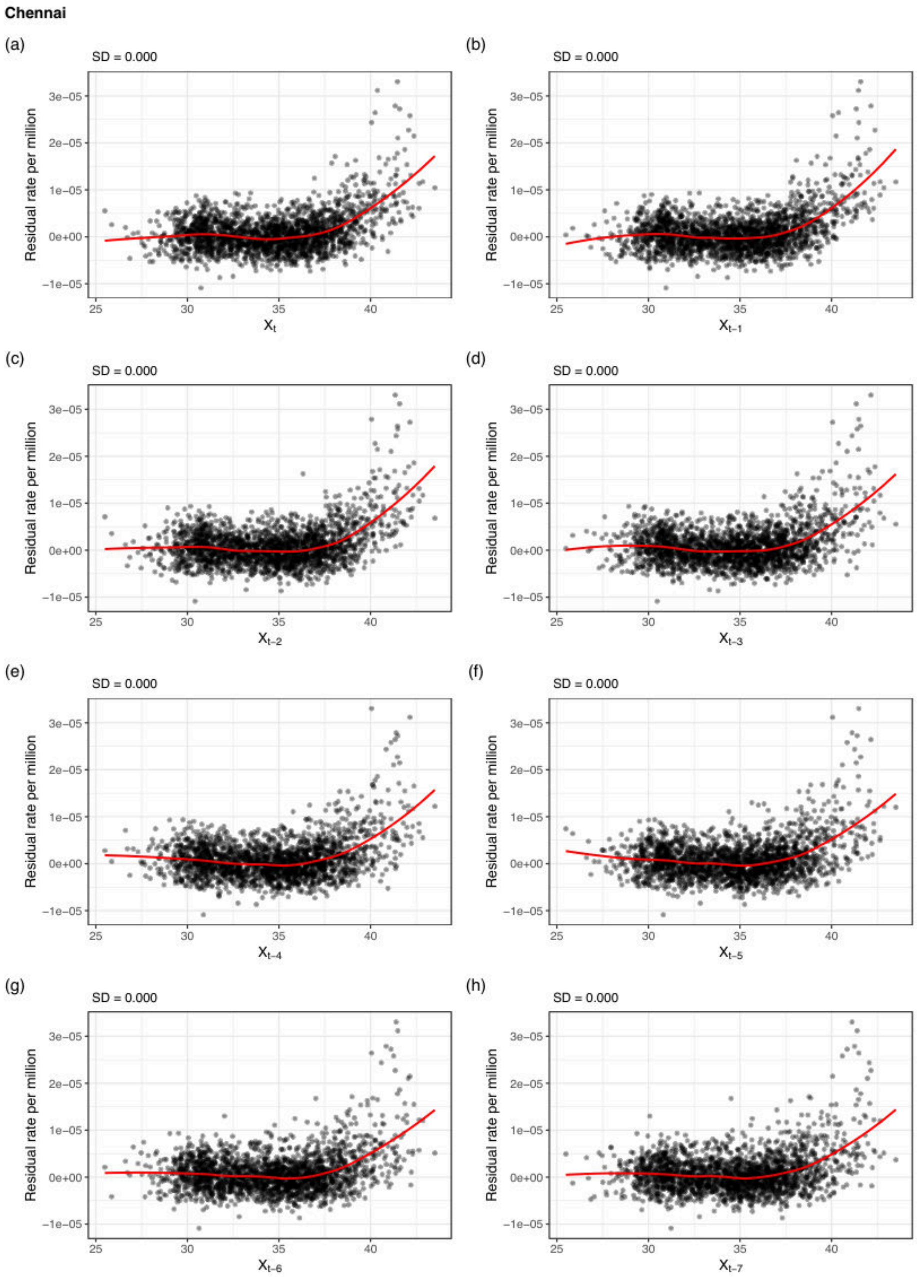


Supplementary Figure S10: Chennai: Long-term fit residuals rate per million versus temperature at each lag. See Figure S9 for full caption.

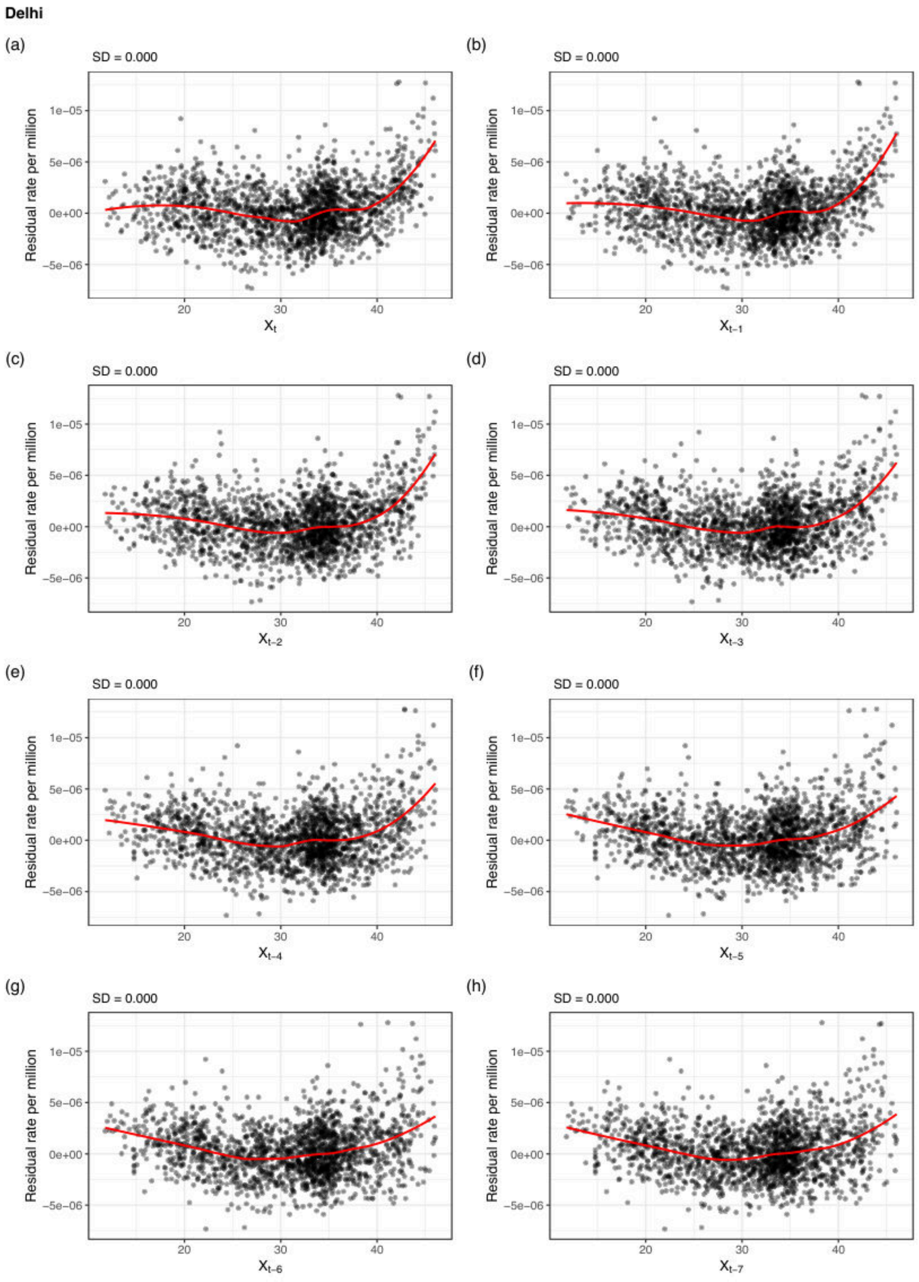


Supplementary Figure S11: Delhi: Long-term fit residuals rate per million versus temperature at each lag. See Figure S9 for full caption.

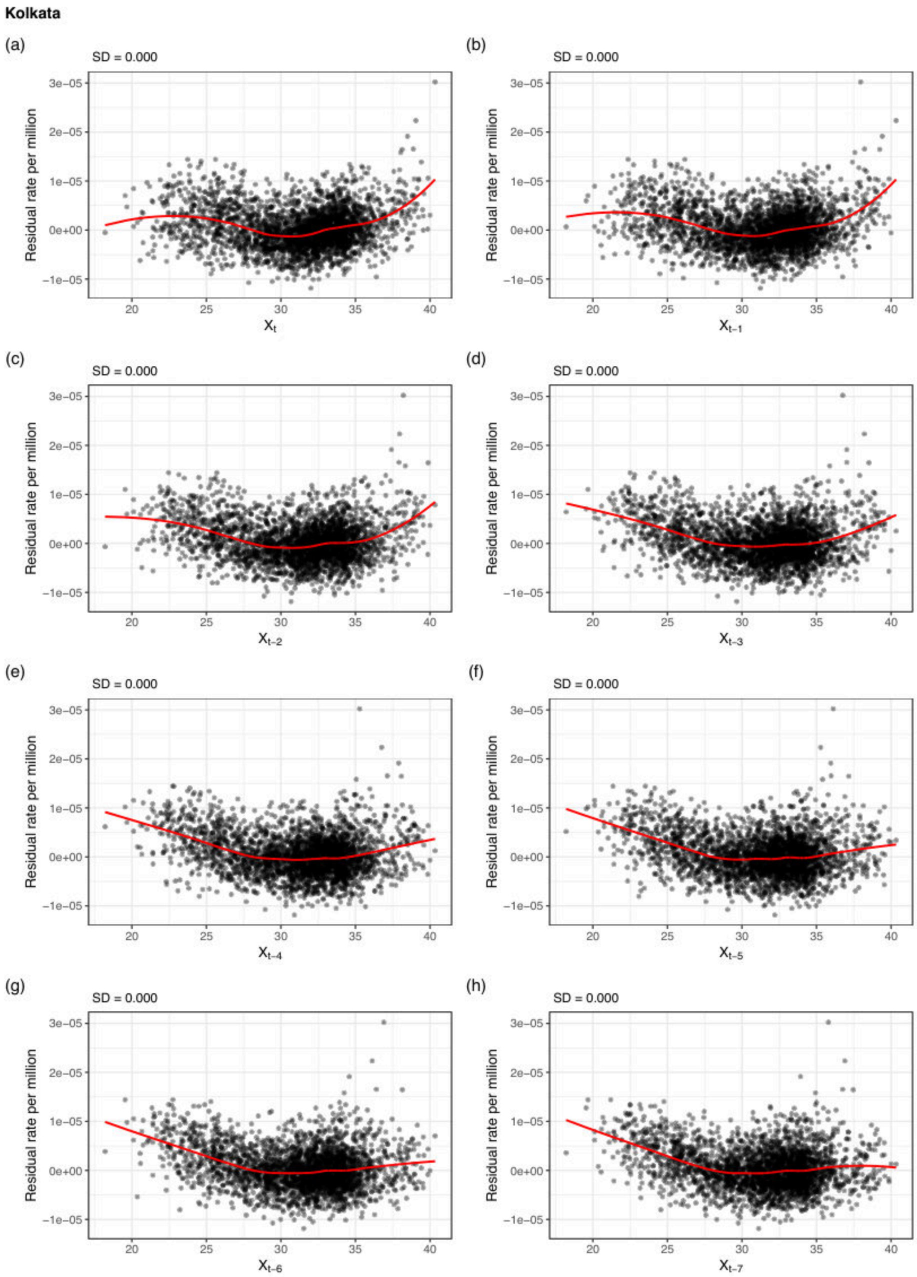


Supplementary Figure S12: Kolkata: Long-term fit residuals rate per million versus temperature at each lag. See Figure S9 for full caption.

**Mumbai**

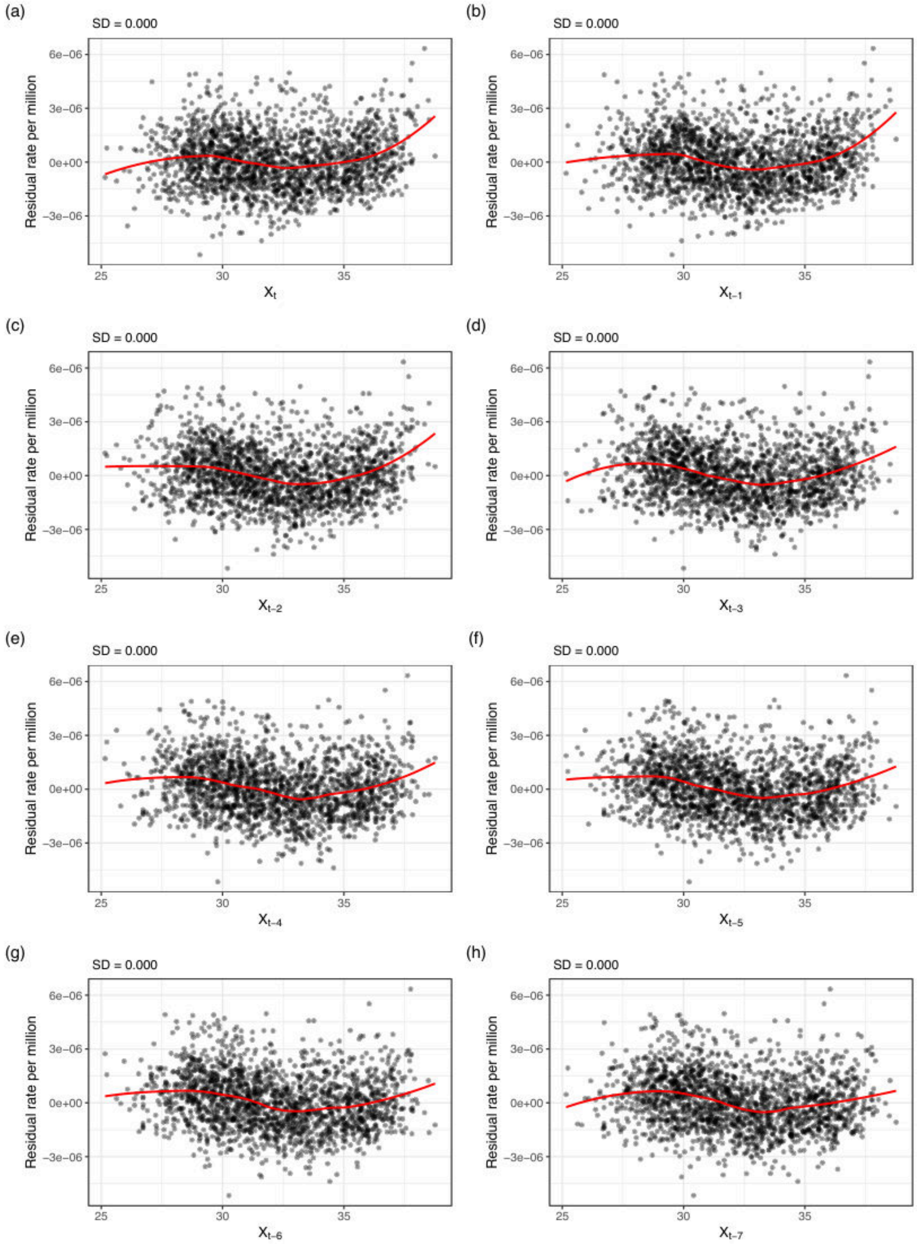


Supplementary Figure S13: Mumbai: Long-term fit residuals rate per million versus temperature at each lag. See Figure S9 for full caption.

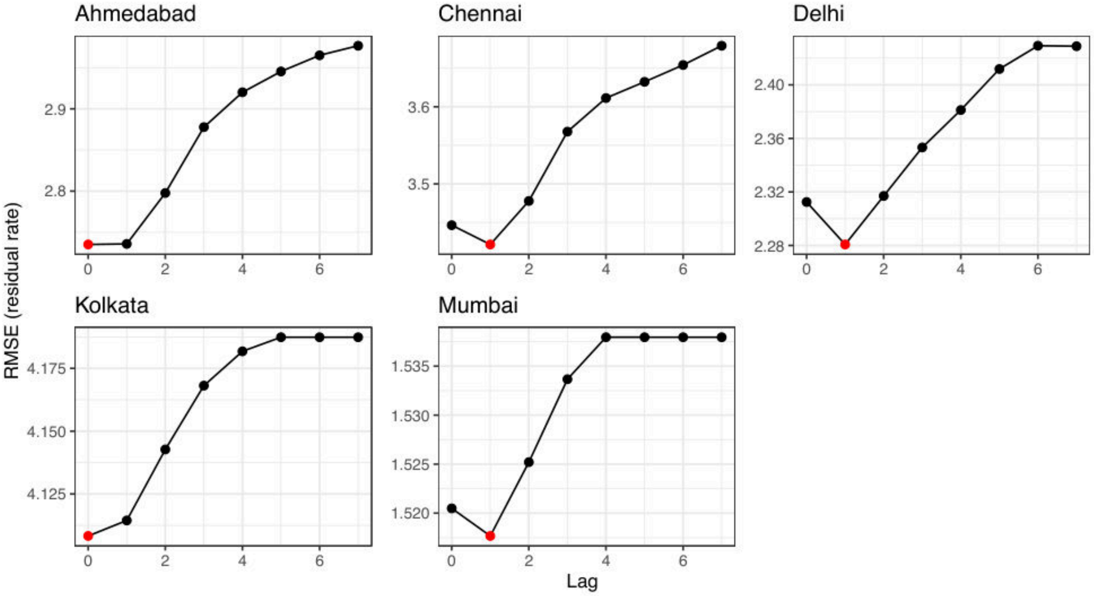


Supplementary Figure S14: Root mean squared error (RMSE) of residual mortality rate (per million) for each model across five Indian cities. For each city, we fit the proposed structured model using temperature at time $t - l$, for $l \in \{0, 1, 2, 3, 4, 5, 6, 7\}$. We then compute the RMSE of the residuals from each model. Red points indicate the lag at which RMSE is minimized. Across all cities, RMSE is minimized at lag 0 (same-day temperature) or lag 1 (previous-day temperature).

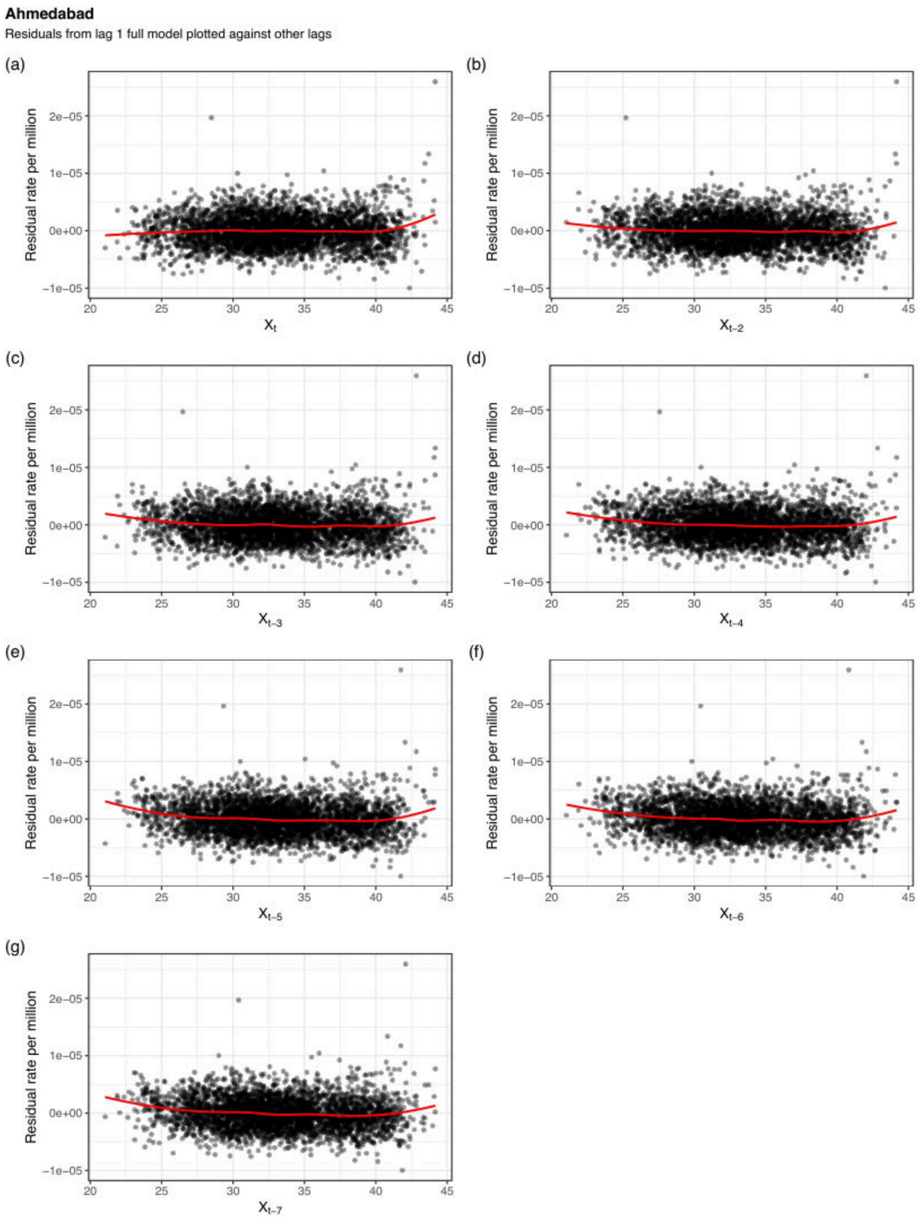


Supplementary Figure S15: Ahmedabad: Full model residual diagnostics after fitting a lag 1 model against temperatures at other lags. For each city, we fit the proposed structured threshold model using temperature at time $t-1$, and compute residual mortality rates (per million person-days) from this model. We then plot these residuals against temperatures at other lags $t-l$, for $l \in \{0, 2, 3, 4, 5, 6, 7\}$. For each lag, we fit a univariate LOESS smoother to model residual mortality rate as a function of temperature, using a local linear fit (degree = 1) with span = 0.5. Panels (a)–(g) show lag-specific residuals and corresponding LOESS smooths. The resulting fitted curves are approximately flat, indicating little remaining association between mortality and temperatures at other lags after accounting for temperature at lag 1.

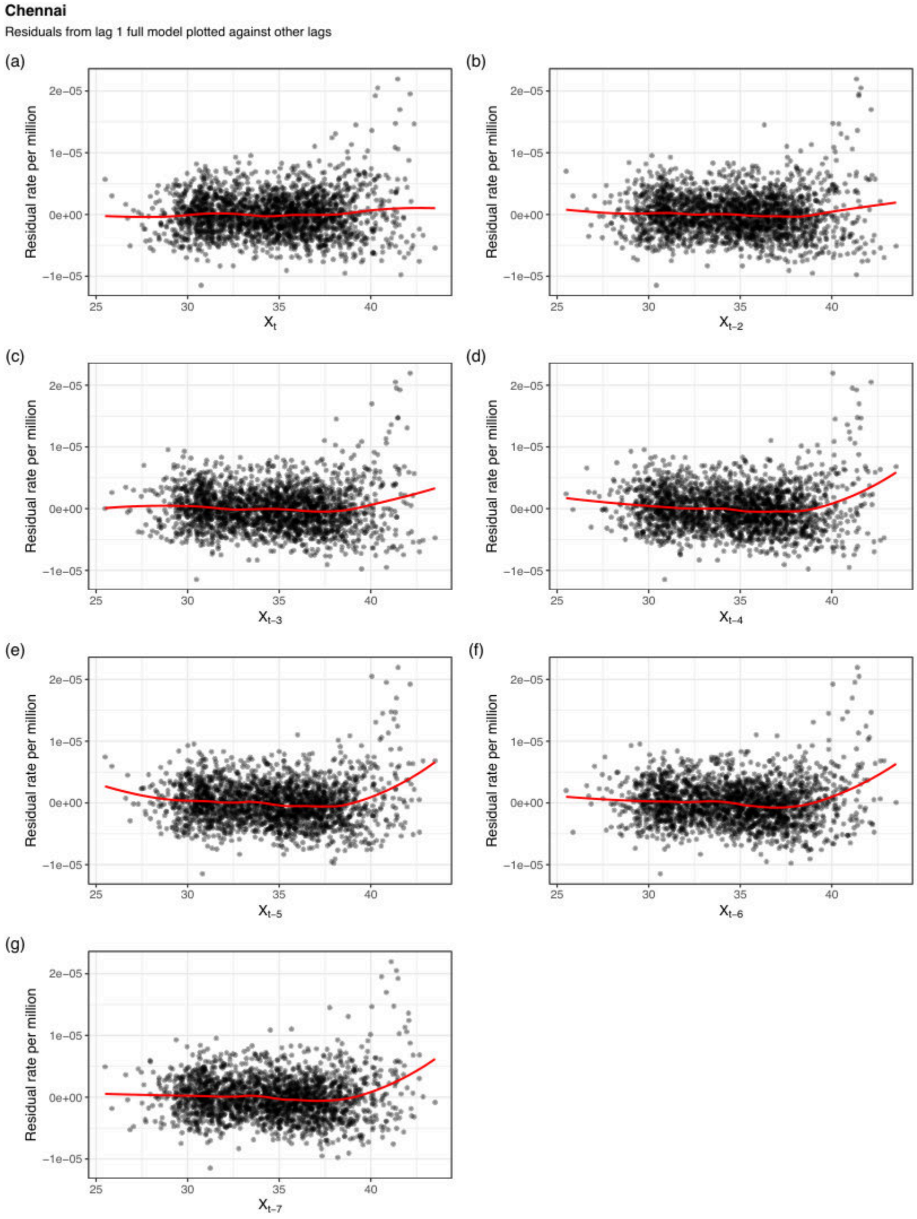


Supplementary Figure S16: Chennai: Full model residual diagnostics after fitting a lag 1 model against temperatures at other lags. See Figure S15 for full caption.

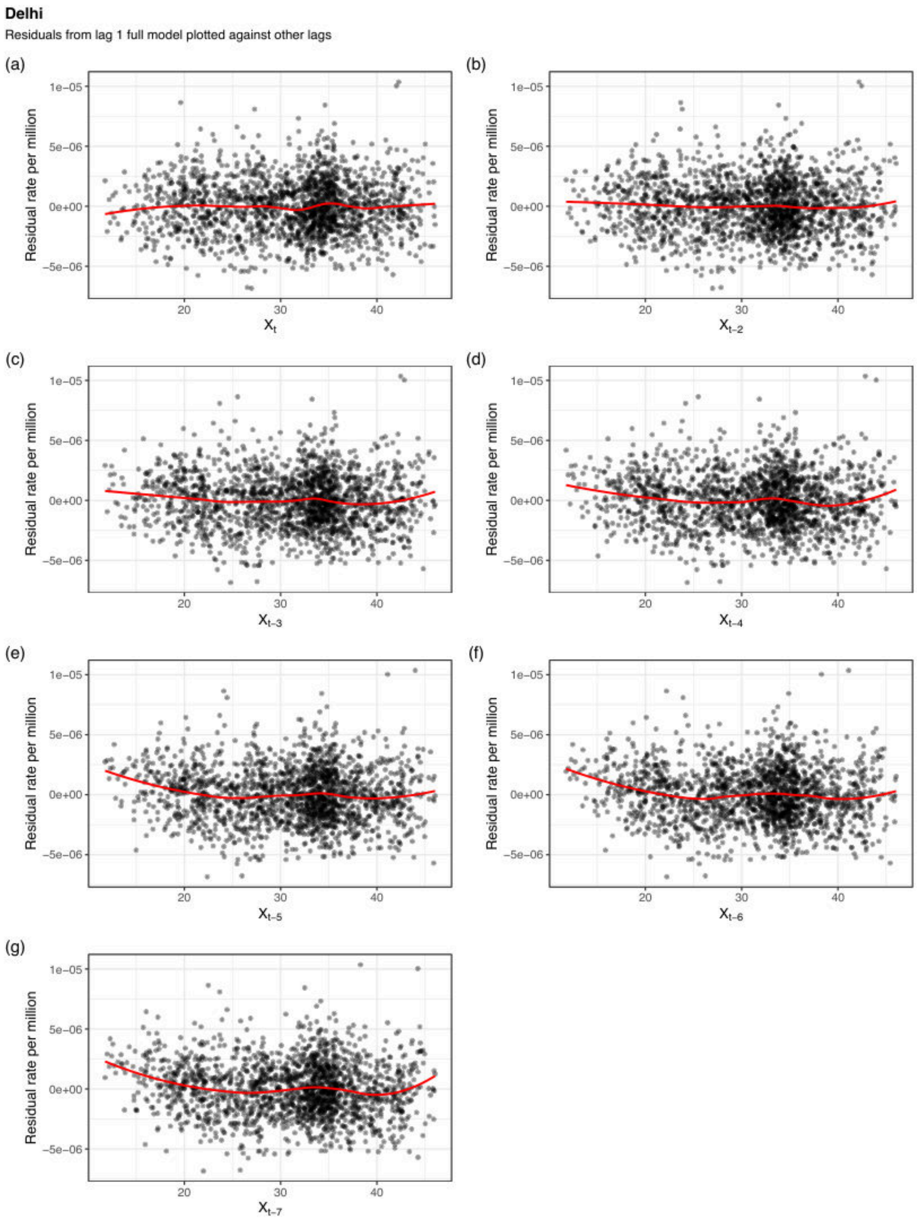


Supplementary Figure S17: Delhi: Full model residual diagnostics after fitting a lag 1 model against temperatures at other lags. See Figure S15 for full caption.

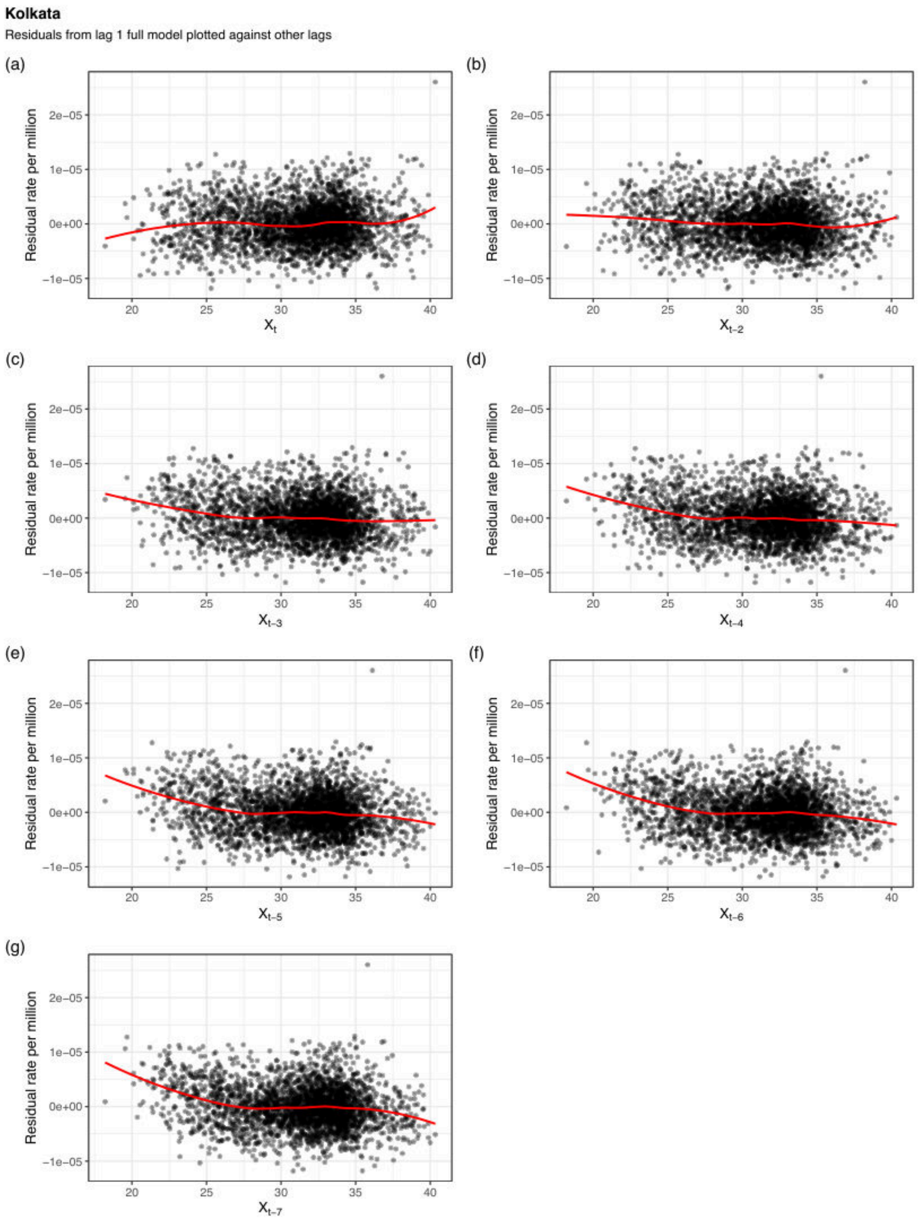


Supplementary Figure S18: Kolkata: Full model residual diagnostics after fitting a lag 1 model against temperatures at other lags. See Figure S15 for full caption.

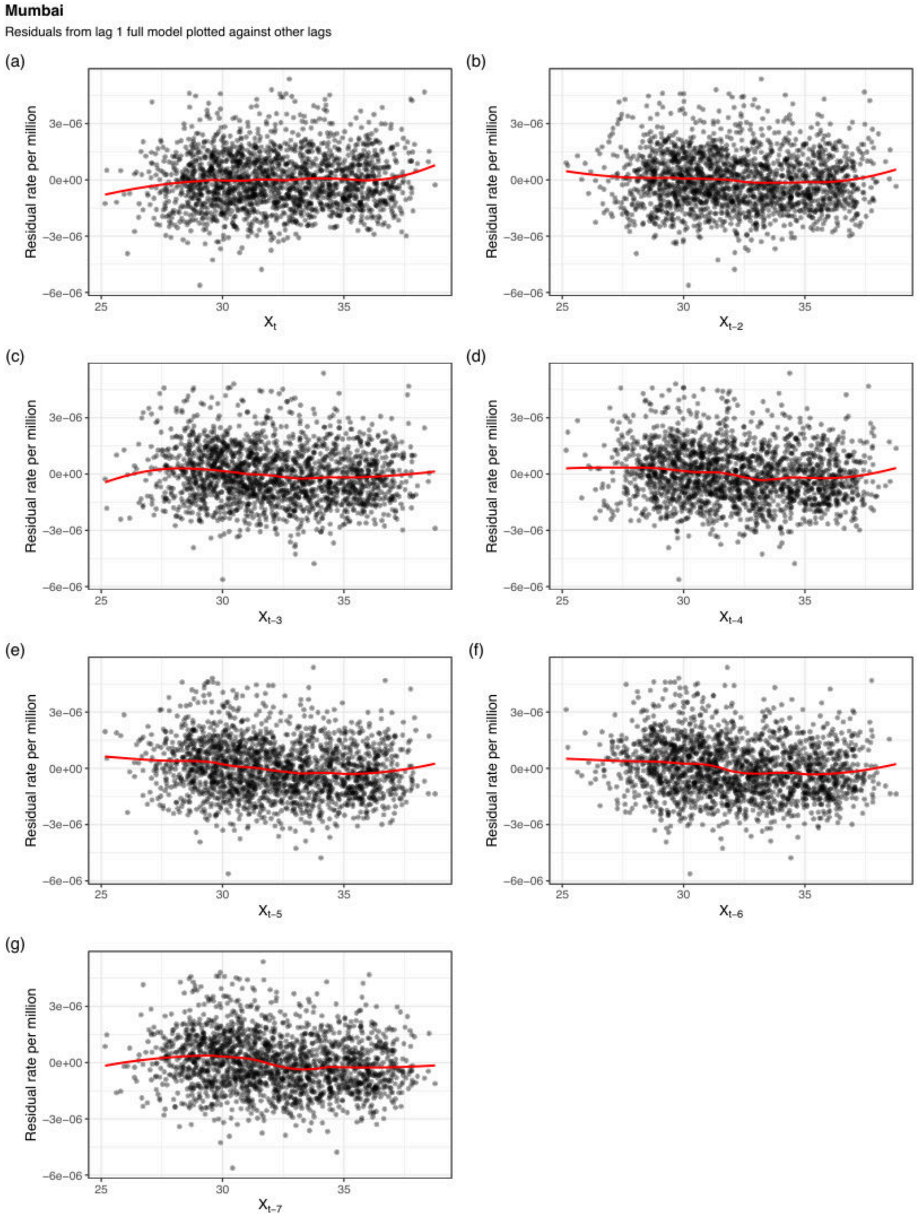


Supplementary Figure S19: Mumbai: Full model residual diagnostics after fitting a lag 1 model against temperatures at other lags. See Figure S15 for full caption.

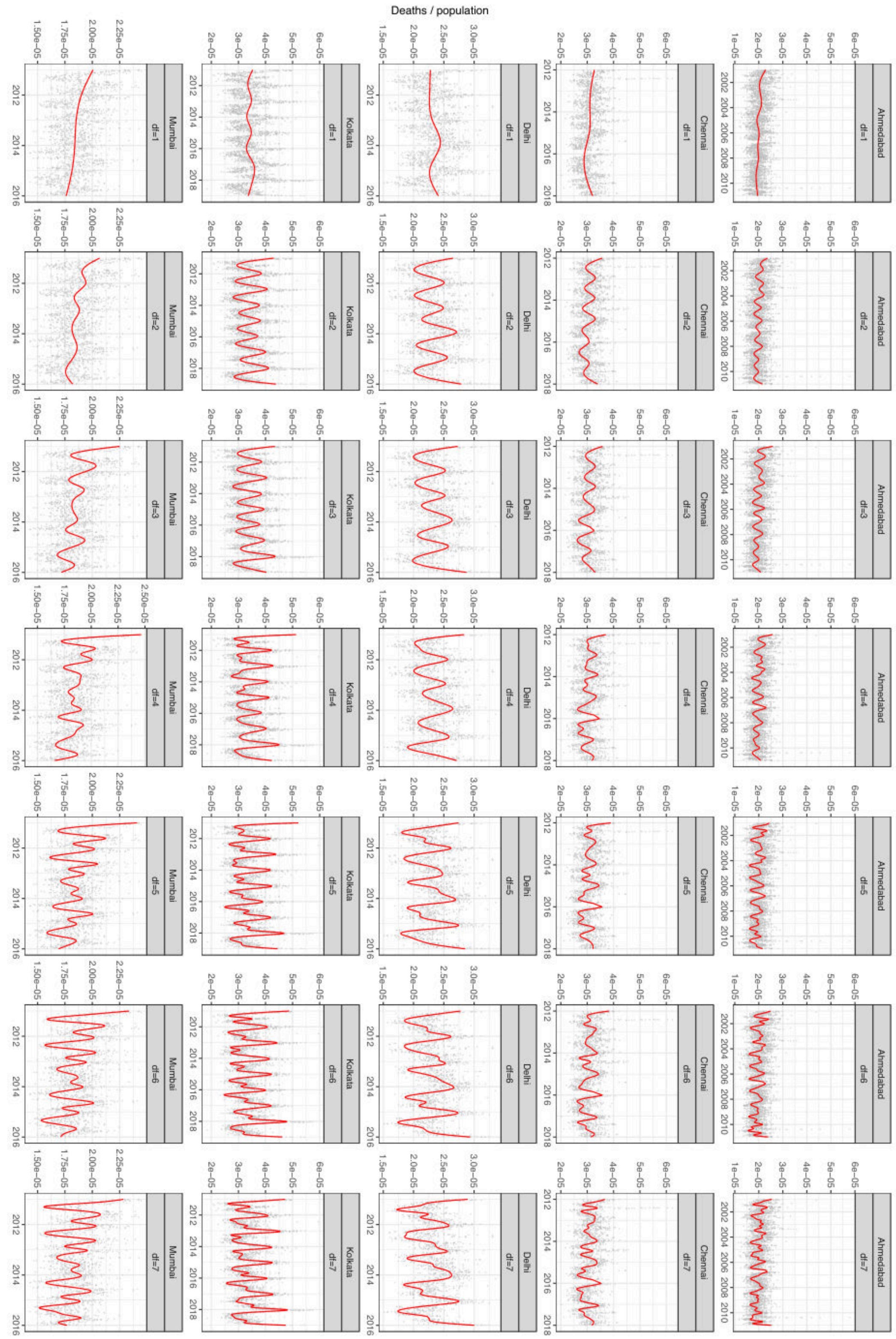


Supplementary Figure S20: Long-term trends with varying degrees of freedom per year for five Indian cities. For each city we fit models with long-term and seasonal trends modeled with natural cubic splines with 1, 2, 3, 4, 5, 6, and 7 degrees of freedom per year of data. The gray points represent the observed mortality counts while the solid red curve represents fitted long-term and seasonal trend. the Degrees of freedom over 4 and 5 capture some of the heat events that inform the temperature–mortality risk curve.

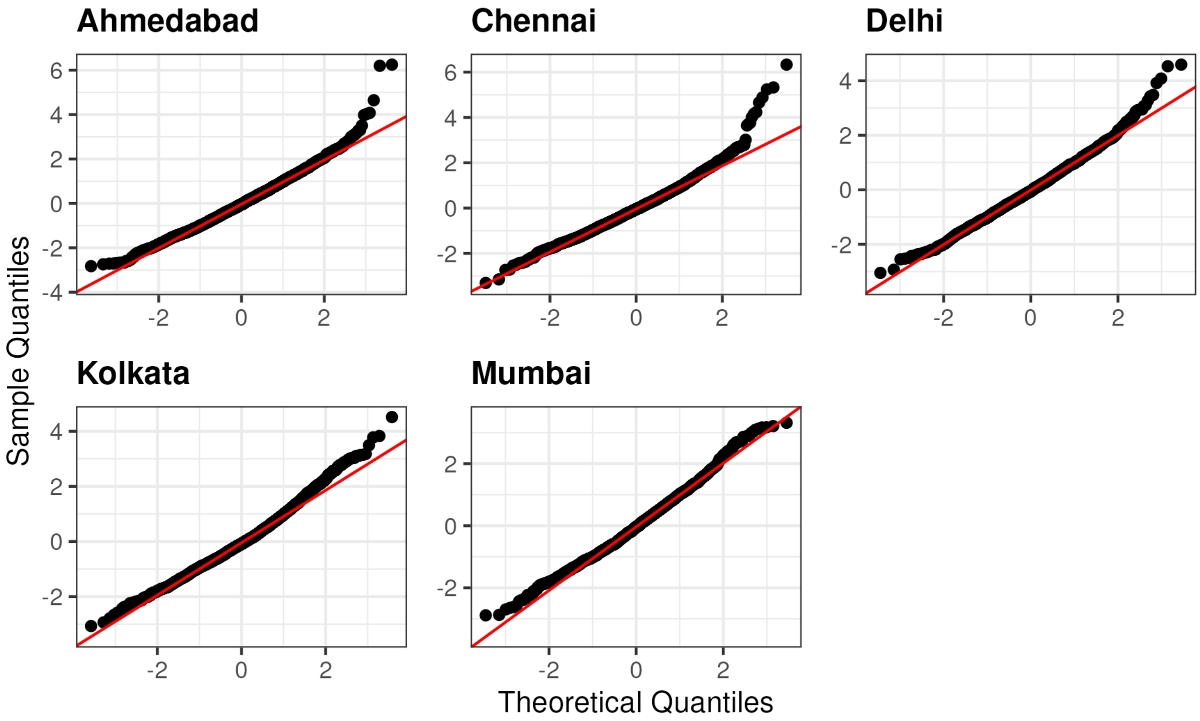


Supplementary Figure S21: Q–Q plots of Pearson residuals from the structured threshold model for each city. Points closely follow the reference line across most of the distribution, indicating adequate model fit. Deviations in the upper tails reflect a small number of observations with elevated mortality, consistent to extreme heat events.
